# Hospital staff views on the visibility, role and impact of Acute Learning Disability Liaison Services in Wales: a service evaluation

**DOI:** 10.64898/2026.06.16.26355793

**Authors:** Abubakar Sha’aban, Francesca Mazzaschi, Ala’a Alazizi, Melanie McAulay, Adrian Edwards, Natalie Joseph-Williams

## Abstract

People with a learning disability experience marked health inequalities. In Wales, Acute Learning Disability Liaison Services (ALDLS) are delivered by specialised learning disability services, and all roles within them are undertaken by Learning Disability Liaison Nurses (LDLN). These services aim to enable access to, and delivery of, secondary care by supporting reasonable adjustments, facilitating communication, and coordinating care for people with learning disability during hospital encounters. However, independent evidence of the impact of ALDLS on patient care remains limited. This evaluation tries to address this evidence gap by examining hospital staff perceptions of the visibility, role, and impact of ALDLS across Welsh Health Boards, with the aim of informing service design and development and improving secondary care access and care for people with learning disability.

The service evaluation used a qualitative approach involving interviews and a focus group with hospital staff across the seven Welsh Health Boards who had experience working with or interacting with ALDLS staff to care for patients with learning disability.

Findings cover six key areas including i) visibility and delivery of ALDLS, ii) Barriers and challenges to effective ALDLS delivery, iii) Enablers of effective ALDLS delivery, iv) Positive impacts for patients with learning disability, v) Negative impacts and unintended consequences when the service is absent or limited, and vi) Participants’ recommendations for future improvements of ALDLS.

To synthesise the findings, we developed an overview diagram, which illustrates how ALDLS may influence care quality in acute hospitals. The overview places the liaison service at the centre, showing how organisational enablers and barriers shape its delivery, and how its core functions support improvements in safety, timeliness, effectiveness, efficiency, equity, and patient-centred care.

From the findings we have identified recommendations for practice and policy. These include that ALDLS should be recognised as a core, safety-critical component of acute hospital care for people with a learning disability, rather than an optional add-on. In practice, services should be more visibly embedded within routine pathways, with consistent site-based presence, clear referral criteria, early identification through electronic flagging and notification systems, and routine involvement in multidisciplinary planning for complex admissions and procedures. At policy level, ALDLS provision should be recognised within equality and patient safety frameworks as an essential service requiring sustained investment, national minimum configuration standards, adequate staffing, and better-integrated digital systems to support continuity, equitable access, and person-centred care.

## 1 BACKGROUND

People with a learning disability experience significant health inequalities compared to the wider population (NICE, 2021), including earlier avoidable deaths, higher comorbidities, and poorer health outcomes. (Adam Watkins and Rachel Ann Jones, 2024; Kings College London, 2023). In response to these longstanding concerns, ALDLS have been developed across Wales to support equitable and safe access to secondary healthcare. However, provision is not uniform across all Health Boards. Dedicated acute liaison services are established in some areas, while alternative models operate elsewhere. For example, Powys Teaching Health Board (PTHB) provides ALDLS through a primary care liaison service, while Swansea Bay University Health Board (SBUHB) has responsibility for learning disability services across three areas (SBUHB, Cardiff and Vale University Health Board (CAVUHB), and Cwm Taf Morgannwg University Health Board (CTMUHB)) delivering commissioned support across these regions. As a result, the structure, coverage, and delivery of liaison services vary across Wales (Betsi Cadwaladr University Health Board, nd; Cardiff and Vale University Health Board, nd; Cwm Taf Morgannwg University Health Board, nd).Despite these differences, all services share the overarching aim of improving the quality, safety, and equity of care for patients with learning disability in secondary (hospital) care (Welsh Government, 2023).

ALDLS are typically provided by Learning Disability Liaison Nurses (LDLNs), whose roles include ensuring reasonable adjustments for patients with disability, supporting communication, advising clinical teams, and promoting person-centred care throughout the patient’s healthcare journey (Macarthur et al., 2015). Previous research indicates that learning disability liaison services can improve hospital care for people with a learning disability by acting as a bridge between patients, families, and clinical teams, advocating for adjustments, and supporting staff to better understand patients’ needs (Brown et al., 2016; Moloney et al., 2023; Oulton et al., 2022). Studies involving liaison nurses themselves emphasise their role as educators, coordinators, and advocates who helps to integrate specialist learning disability expertise into acute hospital care (Kelleher et al., 2023). Evidence from hospital staff perspectives in paediatric settings, particularly in hospitals with dedicated learning disability nurse provision, suggests improvements in staff confidence, communication, and care coordination (Oulton et al., 2019). However, these findings may not be directly transferable to adult acute services, where service models and patient needs differ substantially. This highlights the need for further evidence within adult acute settings.

A recent evidence synthesis highlighted important gaps in the current knowledge base. The rapid review conducted by the Health and Care Research Wales Evidence Centre (Kiseleva et al., 2025) found that much of the published literature on learning disability liaison services has been authored by researchers affiliated with the services being evaluated, including liaison nurses themselves. While these studies provide valuable insights into service delivery and perceived benefits, this concentration of insider perspectives introduces a potential risk of bias and underscores the need for more independent evaluations of liaison services.

Addressing this gap is particularly important given the policy and practice implications associated with liaison services, which are often positioned as key mechanisms for reducing health inequalities for people with a learning disability within hospital settings. Independent research examining how these services are perceived and experienced by the wider hospital workforce can therefore provide a more balanced understanding of their operational impact and implementation challenges.

The success of ALDLSs depends not only on their design and outcomes but also on how they are understood, accessed, and utilised by the wider hospital workforce. Hospital staff (including those working in emergency departments, inpatient wards, and outpatient services) play a central role in identifying patients with a learning disability, initiating referrals, and implementing recommended adjustments in day-to-day care. Their awareness of the service, confidence in its role, and experiences of working alongside liaison nurses delivering the service are therefore critical to whether the service is accessed and integrated into routine care. However, little is currently known about how ALDLS are perceived and experienced by secondary care (hospital) staff in Wales, where service models vary across Health Boards.

This service evaluation aims to address the identified evidence gap by providing an independent examination of hospital staff experiences of ALDLS across Welsh Health Boards. The evaluation focuses exclusively on the experiences and perspectives of hospital staff who worked with or referred patients with a learning disability to ALDLS Teams in secondary care hospital-based settings across Wales.

### 1.1 Aims and Objectives

#### 1.1.1 Aim

To explore hospital staff experiences and perceptions of the visibility, role, and impact of the ALDLS across Welsh Health Boards, to understand how the service supports the delivery of equitable, safe, and person-centred care for patients with a learning disability in secondary care settings.

#### 1.1.2 Objectives

1. To explore how hospital staff understand the purpose of the ALDLS and their experiences of ALDLS delivery and interacting with the service
2. To explore the perceived impact of the ALDLS across the six domains of health care quality (Institute of Medicine Committee on Quality of Health Care in America, 2001; NHS Wales, 2022):

- Safety – whether the service contributes to reducing harm or risks.
- Timeliness – whether it helps avoid delays in health care.
- Effectiveness – whether it contributes to improved patient outcomes or processes.
- Efficiency – whether it supports streamlined, coordinated health care.
- Equity – whether it improves fair access and support for patients with a learning disability.
- Patient-centredness – whether it enhances individualised, respectful care.
3. To assess how well the ALDLS is implemented in practice, including:

- Availability and accessibility of the service.
- Integration into routine workflows and health care processes.
- Adaptations, barriers, and facilitators to effective implementation.
4. To identify examples of good practice and challenges, including:

- Specific instances of positive or negative impact.
- Hospital staff confidence in supporting patients with a learning disability.
5. To gather hospital staff recommendations for improvement and sustainability of the ALDLS:

- Enhancements needed for greater impact of ALDLS on LD patient care.
- Suggestions for scale, spread, or adaptation across departments or health boards.

## 2 METHODS

### 2.1 Project setting and population

The project was conducted across the seven Health Boards in Wales: Aneurin Bevan University Health Board (ABUHB); Betsi Cadwaladr University Health Board (BCUHB); Cardiff and Vale University Health Board (CAVUHB); Cwm Taf Morgannwg University Health Board (CTMUHB); Hywel Dda University Health Board (HDUHB); Powys Teaching Health Board (PTHB); and Swansea Bay University Health Board (SWUHB). ALDLS are organised variably across these Health Boards. Four Health Boards (ABUHB, BCUHB, HDUHB, and PTHB) each provide their own ALDLS. In contrast, Swansea Bay University Health Board is commissioned to deliver ALDLS both within its own Health Board and on behalf of two neighbouring Health Boards (CAVUHB and CTMUHB), reflecting a shared service delivery model in parts of Wales. Additionally, unlike the other six Health Boards, Powys Teaching Health Board provides ALDLS through a primary care liaison model rather than a secondary care-based service.

The project population includes hospital staff (e.g., nurses, accident and emergency (A&E) department staff, ward staff, and outpatient clinicians) who have experience working with or referring to ALDLS in hospital environments.

### 2.2 Project design

This service evaluation adopted a qualitative design, using interviews and a focus group, to explore the views and experiences of hospital staff regarding the ALDLS across the seven Health Boards in Wales. The evaluation is underpinned by two key frameworks:

**The six domains of health care quality** (Institute of Medicine Committee on Quality of Health Care in America, 2001; NHS Wales, 2022) to assess perceived impact of the service in terms of safety, timeliness, effectiveness, efficiency, equity, and patient-centredness.

**The MRC Process Evaluation Framework** (Graham Moore, 2015) (see Figure 1) to examine how the service is implemented, how it interacts with context, and how it leads to outcomes.

**Figure 1.**
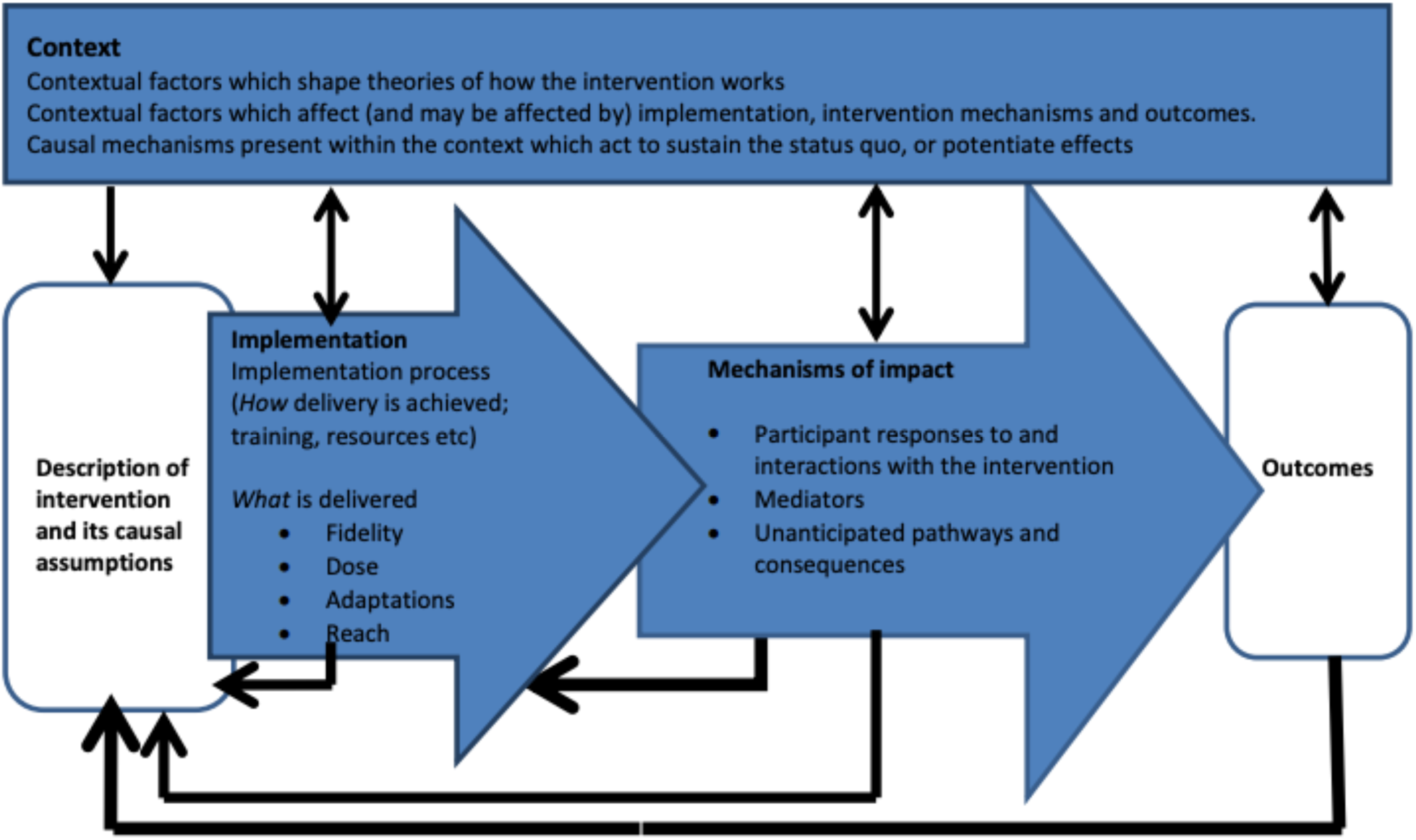
MRC Process Evaluation Framework.

This dual framework enables both a structured understanding of health care quality, and in-depth exploration of how ALDLS function in real-world secondary care settings and adaptations made in practice to try to enhance patient, family or staff experiences. This dual framework helped in designing the interview and focus group guides for this service evaluation.

#### 2.2.1 Six domains of health care quality

The six domains of health care quality are an operational framework for quality improvement set out by the Institute of Medicine (2001). Quality is defined as meeting everyone’s health care needs consistently every time and all the time. The six domains of quality are commonly referred to using the acronym STEEEP. In our context, the six domains of health care quality provide a lens through which hospital staff can evaluate the impact of the ALDLS on care delivery. Interview questions explored:

- **Safety**-Whether the service helps avoid harm
- **Timeliness-**Whether it contributes to reducing delays
- **Effectiveness**-Whether it improves outcomes
- **Efficiency-**Whether it supports efficient use of resources
- **Equity-**Whether it improves access and outcomes equitably
- **Patient-centredness-** Whether it enables person-centred care and better communication

#### 2.2.2 MRC Process Evaluation Framework

The Medical Research Council (MRC) describes the importance of process evaluation in understanding an intervention and its results by developing an understanding of “fidelity and quality of *implementation*, clarifying causal *mechanisms* and identifying *contextual factors* associated with variation in outcomes” (Graham Moore, 2015). These key themes of context, mechanisms and implementation have dynamic relationships as illustrated in **Figure 1** (below) from the MRC. Contextual factors are from outside the intervention but can affect it, such as a barrier to someone carrying out their role. Implementation is a process of how the intervention is delivered and includes fidelity (how well people are able to carry out the role according to the original aims), dose (whether a patient has had the full care that would be planned), adaptations (changes to the original planned role which may be seen as improvements or problem-solving) and reach (how many of the right patients are being seen). Mechanisms of impact describes the participant responses to the intervention (how patients respond to their care), mediators and unanticipated responses. Together these factors lead to the intervention outcomes.

### 2.3 Participant recruitment

#### 2.3.1 Interview recruitment

Eligible participants were hospital staff who had utilised the ALDLS, either through formal LD patient referral or other professional engagement with the service.

Recruitment was coordinated through Acute Learning Disability Liaison Nurses (LDLNs) within each participating Health Board, identified via project stakeholders. LDLNs disseminated invitation emails outlining the purpose of the evaluation and participation requirements through their referral and professional contact networks. An electronic recruitment poster (Appendix 1) and participant information sheet (Appendix 2) were included, and recipients were encouraged to share the invitation within their teams to facilitate wider dissemination (snowball sampling).

Interested individuals registered their interest by scanning a QR code on the recruitment poster or via a direct link included in the email, which directed them to a Microsoft Forms screening and consent form. Information collected through the form was used to support purposive sampling and ensure diversity of perspectives. Sampling aimed to achieve representation across:

- the seven Welsh Health Boards
- different secondary care contexts (e.g. inpatient, outpatient, emergency settings)
- staff roles (e.g. nurses, allied health professionals, consultants)
- varying levels of engagement with the ALDLS (frequent and infrequent users)
- different shift patterns (e.g. weekday and 24/7 services)

Individuals selected for interview were subsequently contacted by a member of the research team to confirm eligibility and arrange an interview at a mutually convenient time. We aimed to recruit approximately 20–30 participants, guided by principles of information power and data saturation.

#### 2.3.2 Focus group recruitment

Following preliminary analysis of interview data, we identified that most respondents were in managerial roles, resulting in limited representation from frontline, non-managerial staff (e.g. Band 5/6 nurses). A targeted follow-up focus group was therefore designed to address this gap.

Focus group recruitment targeted frontline, patient-facing, and non-managerial hospital staff across Welsh Health Boards. An invitation email outlining the purpose of the evaluation and participation requirements was disseminated via:

- Interview participants
- LDLNs
- Senior managers across Health Boards

An electronic recruitment poster (Appendix 4) was attached, and recipients were encouraged to share the invitation within their teams (snowball dissemination).

Interested individuals registered via Microsoft Forms and were subsequently contacted by a member of the research team to confirm eligibility and allocate a session date.

### 2.4 Data Collection

#### 2.4.1 Interview data collection

Semi-structured interviews were conducted online via Microsoft Teams and lasted approximately 20–60 minutes, depending on participant availability. Interviews were scheduled at mutually convenient times to accommodate clinical commitments.

Prior to interview:

- Participants received a Participant Information Sheet detailing the evaluation and data handling procedures.
- Online informed consent was obtained via Microsoft Forms.
- Interview arrangements were confirmed by email.

Interviews were recorded and automatically transcribed using MS Teams. Transcripts were cross-checked for accuracy, anonymised, and imported into NVivo version 15 (Lumivero, 2025) for analysis. Interviews were guided by a semi-structured interview guide (Appendix 5), informed by the Institute of Medicine’s six domains of health care quality and the Medical Research Council (MRC) process evaluation framework. The guide was co-developed with the project stakeholder group and the project patient and public involvement (PPI) member (MMc). The guide was iteratively refined as analysis progressed. While it provided a structured framework, interviews remained flexible to allow exploration of participant experiences. A summary of key topics explored in presented in Box 1.

**Box 1.**
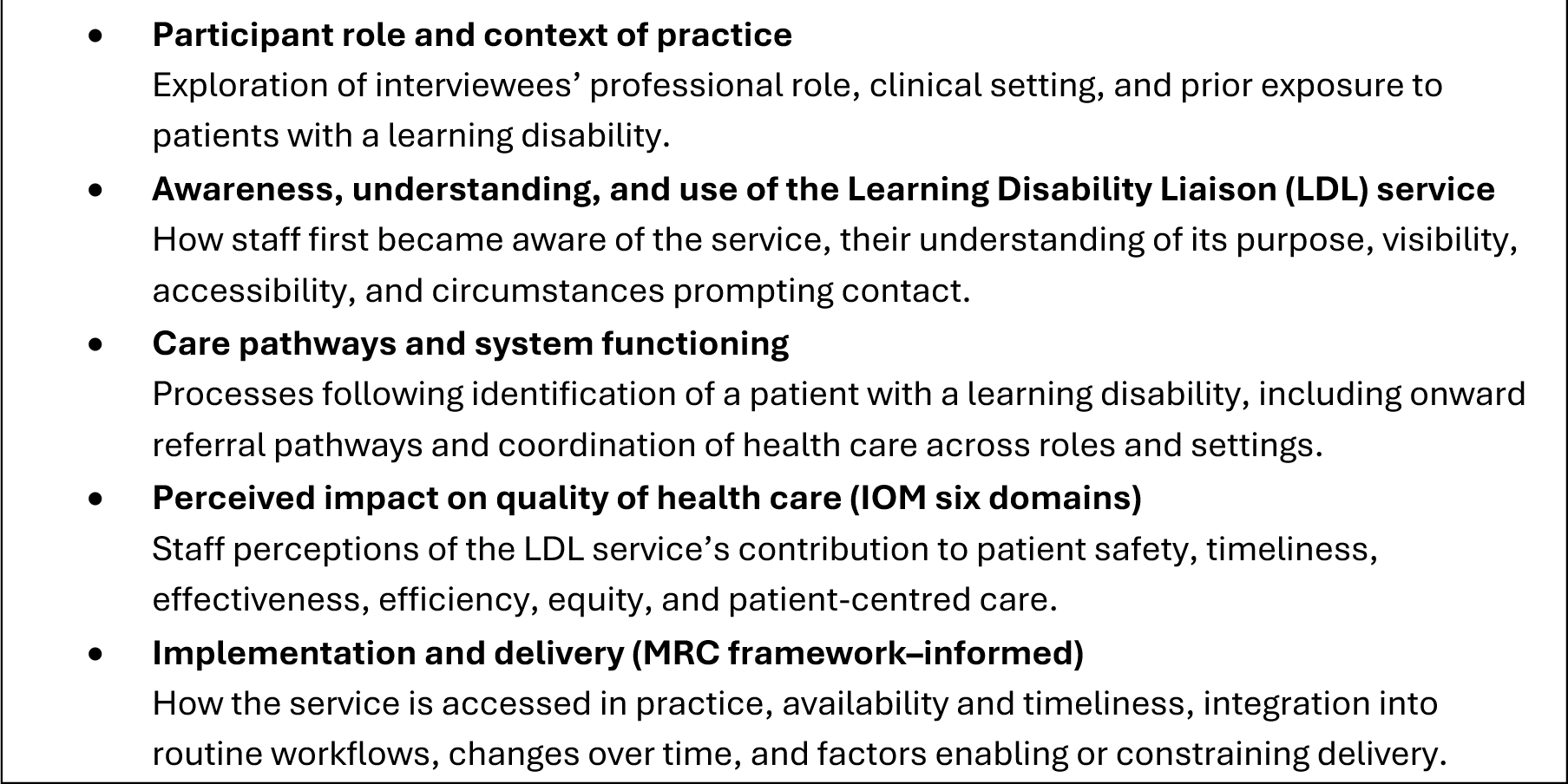
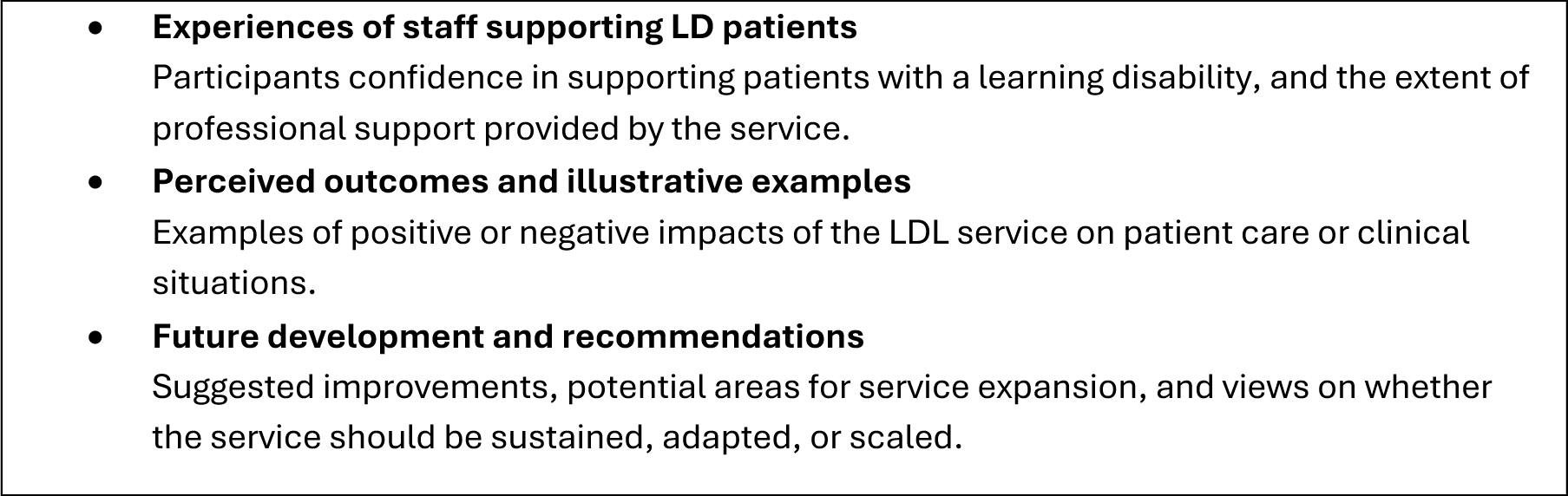
Summary of key topics explored in the Interview schedule.

#### 2.4.2 Focus group data collection

Two online focus group sessions (1 hour each) were scheduled on separate dates to maximise accessibility. The first scheduled session was attended by one participant and was therefore converted into an individual semi-structured interview. The second session proceeded as a focus group, with three participants in attendance (four had registered).

The focus group was conducted using a facilitator guide (Appendix 6) via Microsoft Teams to enable cross-Health Board participation and reduce travel burden. The facilitator guide was designed to:

- Explore experiential perspectives from hospital staff
- Enable comparison with emerging interview themes
- Clarify or extend findings from the earlier interview phase

The focus group was recorded and automatically transcribed using MS Teams. Transcripts were cross-checked for accuracy, anonymised, and uploaded to NVivo version 15 (Lumivero, 2025) for analysis. A summary of key topics explored in the focus group is presented in Box 2.

**Box 2.**
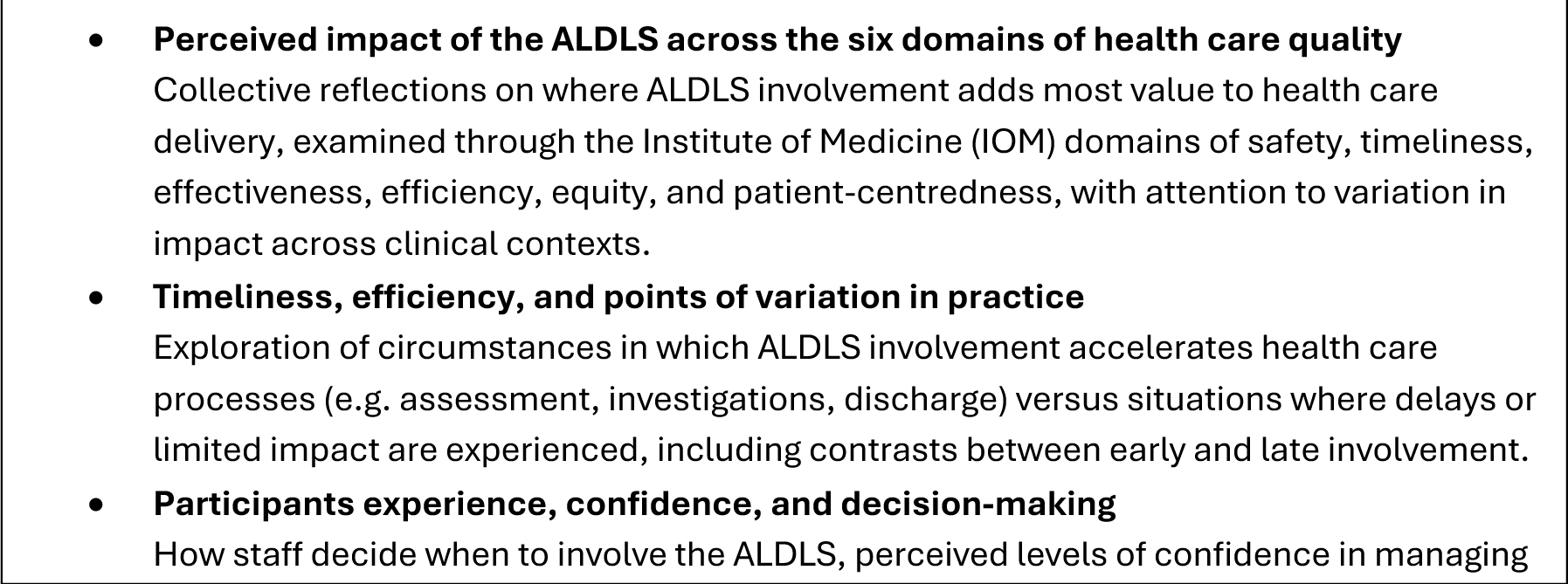
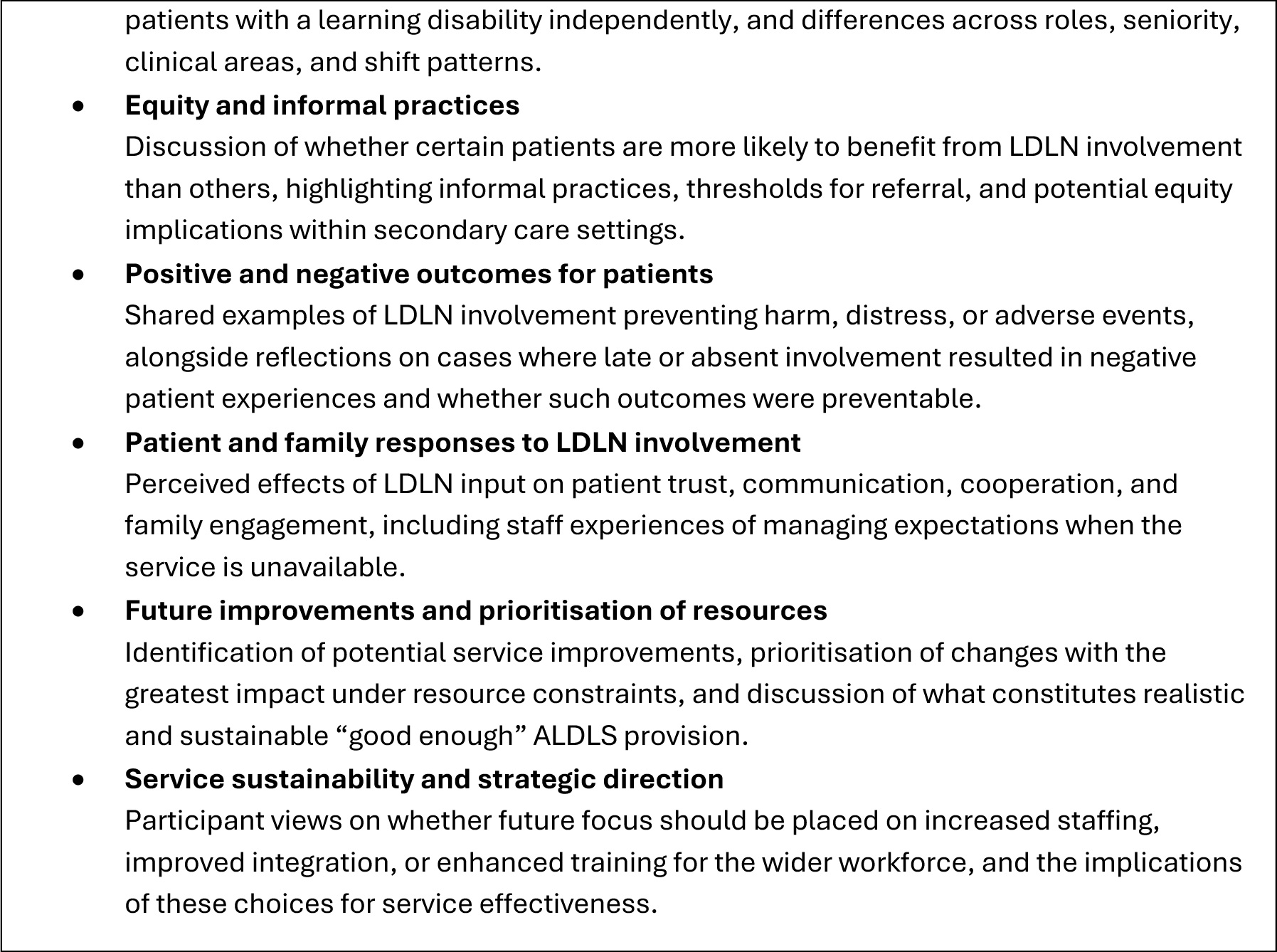
Summary of key topics explored in the focus group guide.

#### 2.4.3 Service-level workforce and organisational data collection

To contextualise staff-reported experiences and better understand variation in service provision across sites, service-level data were collected from each Health Board. The designated Learning Disability Liaison Nurse (LDLN) contact was emailed and asked to provide information on the current number of LDLNs covering acute hospital sites, including any cross-site responsibilities. These data were used descriptively to characterise workforce capacity and organisational configuration across Health Boards, and to support interpretation of findings related to service visibility, accessibility, and reported resource constraints.

### 2.5 Data Analysis

#### 2.5.1 Interview and focus group analysis

Interviews and the focus group transcripts were uploaded into NVivo version 15 (Lumivero, 2025) to facilitate analysis. A framework analysis (Klingberg et al., 2024) approach was used to support systematic coding of the data using both predefined (deductive) and emergent (inductive) themes.

Predefined themes were primarily informed by the six domains of health care quality, with staff accounts coded by domain as evidence of change or impact, and secondly by the Medical Research Council (MRC) Process Evaluation framework (context, implementation, and mechanisms). Inductively generated themes arising from the data were integrated alongside these frameworks.

The analytic process comprised familiarisation with the transcripts, coding aligned to the two frameworks, charting data into a matrix to enable comparison across participants and themes, and interpretative analysis to explore patterns, variations, and implications.

Interviews and the focus group were transcribed verbatim and analysed using NVivo version 15 (Lumivero, 2025). A framework analysis approach was adopted to address the study aims and objectives. An initial coding framework was developed from six interview transcripts and agreed by the research team. This framework was then applied to the remaining transcripts, with additional codes generated inductively where relevant. Emerging themes were reviewed with the wider project team, and the analytic framework was iteratively refined throughout the analysis, with themes merged, reorganised, or further developed as appropriate.

### 2.6 Ethics and governance

This project was classified as a service evaluation using the Health Research Authority (HRA) Decision Tool (https://www.hra-decisiontools.org.uk/research/, developed for the Medical Research Council) and this classification was verified by the Cardiff University Joint Research Office. Permissions to conduct the evaluation were obtained from the Research and Development departments of each Health Board.

All participants provided informed consent via an online Microsoft Forms consent form (Appendix 3) at the point of registration and were provided with a Participant Information Sheet (Appendix 2), which was also emailed to them prior to the interview.

At the start of each interview, and before audio-recording commenced, participants were reminded that their participation was voluntary, that they could withdraw at any time without providing a reason, that they were not required to answer any questions they did not wish to, and that breaks could be taken if needed.

## 3 RESULTS

### 3.1 Study participants and service-level data sources

#### 3.1.1 Interview and Focus group participants

23 participants took part in the intervies and three in the focus group. **Table 1** summarises the demographic and professional characteristics of participants who took part in the interviews and focus group. For each participant, the table presents the study identification number, job role, primary clinical or organisational setting, and length of time working in their current role. These details provide contextual information about the range of professional backgrounds and service settings represented in the study, supporting interpretation of the findings by illustrating the diversity of experience across participants.

**Table 1.**
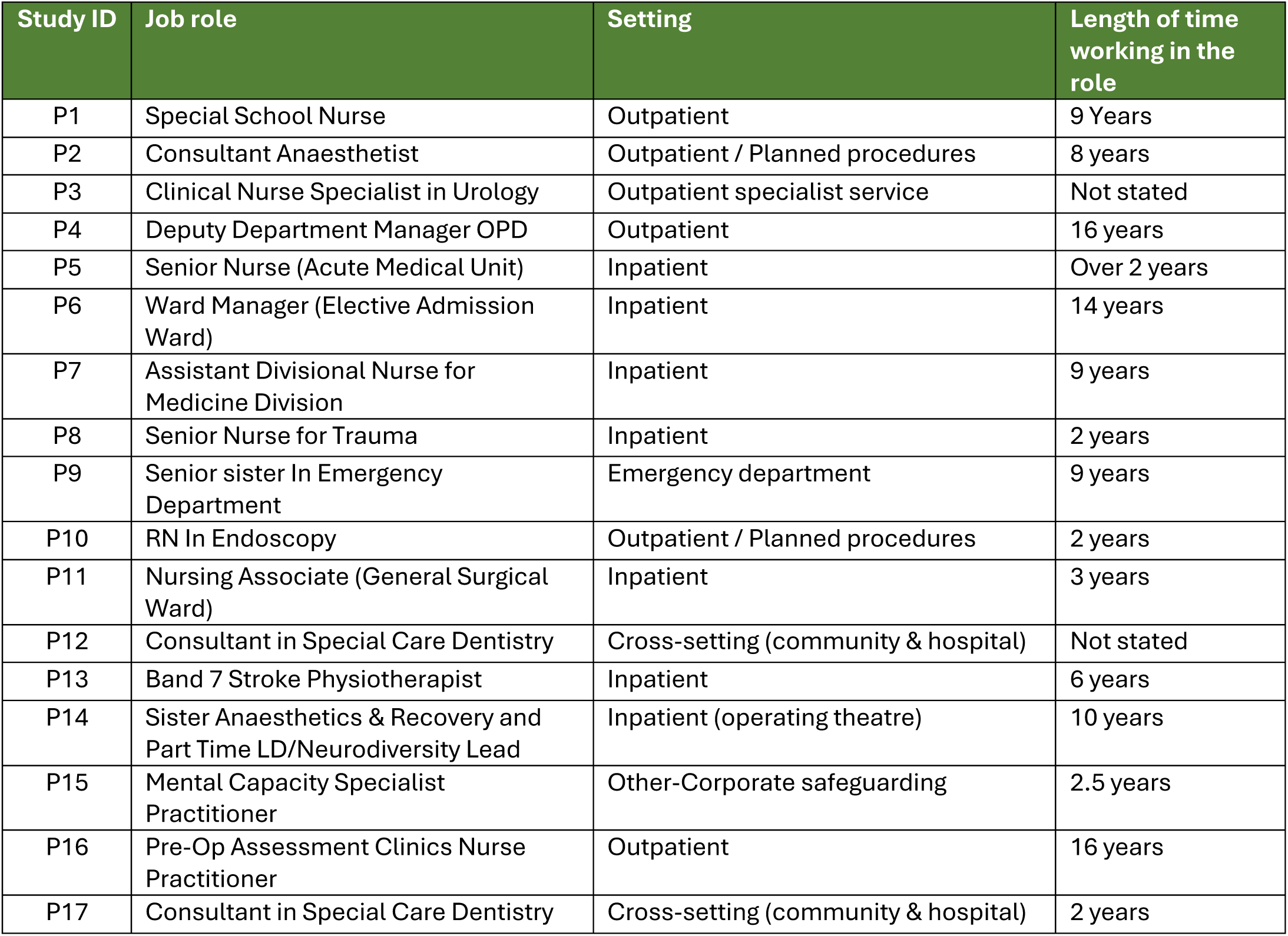
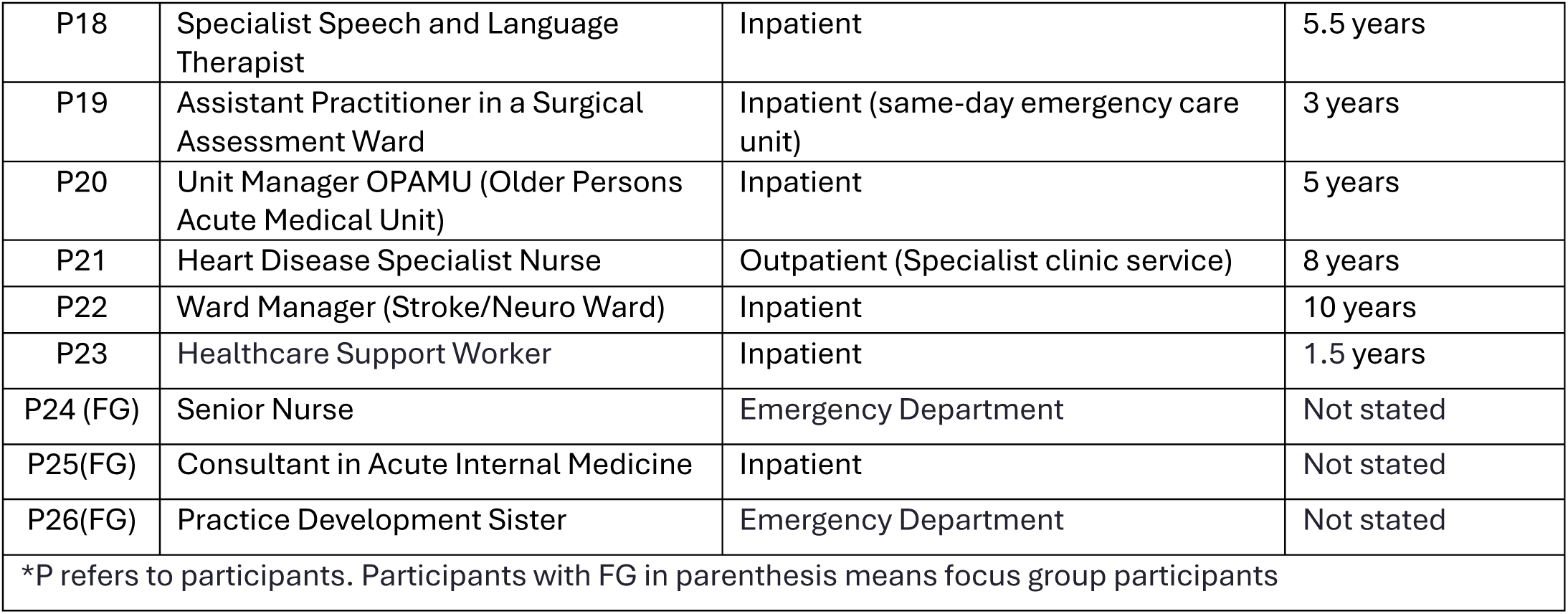
Interview and Focus group demographic data.

| Study ID | Job role | Setting | Length of time working in the role |
| --- | --- | --- | --- |
| P1 | Special School Nurse | Outpatient | 9 Years |
| P2 | Consultant Anaesthetist | Outpatient / Planned procedures | 8 years |
| P3 | Clinical Nurse Specialist in Urology | Outpatient specialist service | Not stated |
| P4 | Deputy Department Manager OPD | Outpatient | 16 years |
| P5 | Senior Nurse (Acute Medical Unit) | Inpatient | Over 2 years |
| P6 | Ward Manager (Elective Admission Ward) | Inpatient | 14 years |
| P7 | Assistant Divisional Nurse for Medicine Division | Inpatient | 9 years |
| P8 | Senior Nurse for Trauma | Inpatient | 2 years |
| P9 | Senior sister In Emergency Department | Emergency department | 9 years |
| P10 | RN In Endoscopy | Outpatient / Planned procedures | 2 years |
| P11 | Nursing Associate (General Surgical Ward) | Inpatient | 3 years |
| P12 | Consultant in Special Care Dentistry | Cross-setting (community & hospital) | Not stated |
| P13 | Band 7 Stroke Physiotherapist | Inpatient | 6 years |
| P14 | Sister Anaesthetics & Recovery and Part Time LD/Neurodiversity Lead | Inpatient (operating theatre) | 10 years |
| P15 | Mental Capacity Specialist Practitioner | Other-Corporate safeguarding | 2.5 years |
| P16 | Pre-Op Assessment Clinics Nurse Practitioner | Outpatient | 16 years |
| P17 | Consultant in Special Care Dentistry | Cross-setting (community & hospital) | 2 years |
| P18 | Specialist Speech and Language Therapist | Inpatient | 5.5 years |
| P19 | Assistant Practitioner in a Surgical Assessment Ward | Inpatient (same-day emergency care unit) | 3 years |
| P20 | Unit Manager OPAMU (Older Persons Acute Medical Unit) | Inpatient | 5 years |
| P21 | Heart Disease Specialist Nurse | Outpatient (Specialist clinic service) | 8 years |
| P22 | Ward Manager (Stroke/Neuro Ward) | Inpatient | 10 years |
| P23 | Healthcare Support Worker | Inpatient | 1.5 years |
| P24 (FG) | Senior Nurse | Emergency Department | Not stated |
| P25(FG) | Consultant in Acute Internal Medicine | Inpatient | Not stated |
| P26(FG) | Practice Development Sister | Emergency Department | Not stated |
| *P refers to participants. Participants with FG in parenthesis means focus group participants |  |  |  |

#### 3.1.2 Service-level data contributors (LDLNs)

Across Health Boards, LDLN staffing levels varied considerably. While some services were relatively well resourced, many comprised small teams covering multiple acute sites and departments, limiting capacity for consistent on-site presence. This variation provides important context for the reported constraints on service visibility, responsiveness, and proactive engagement. Details are presented in **Table 2**.

**Table 2.**
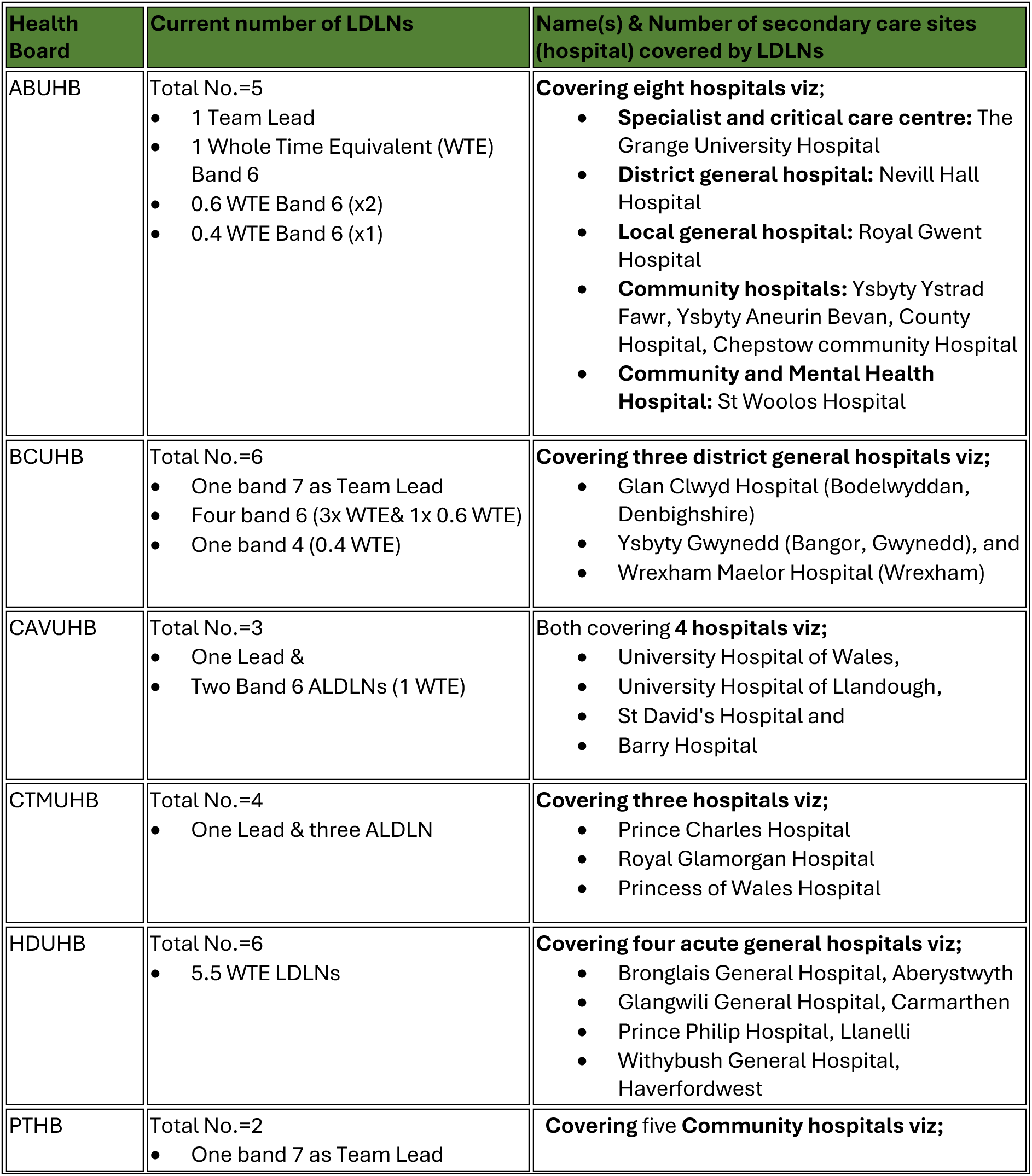
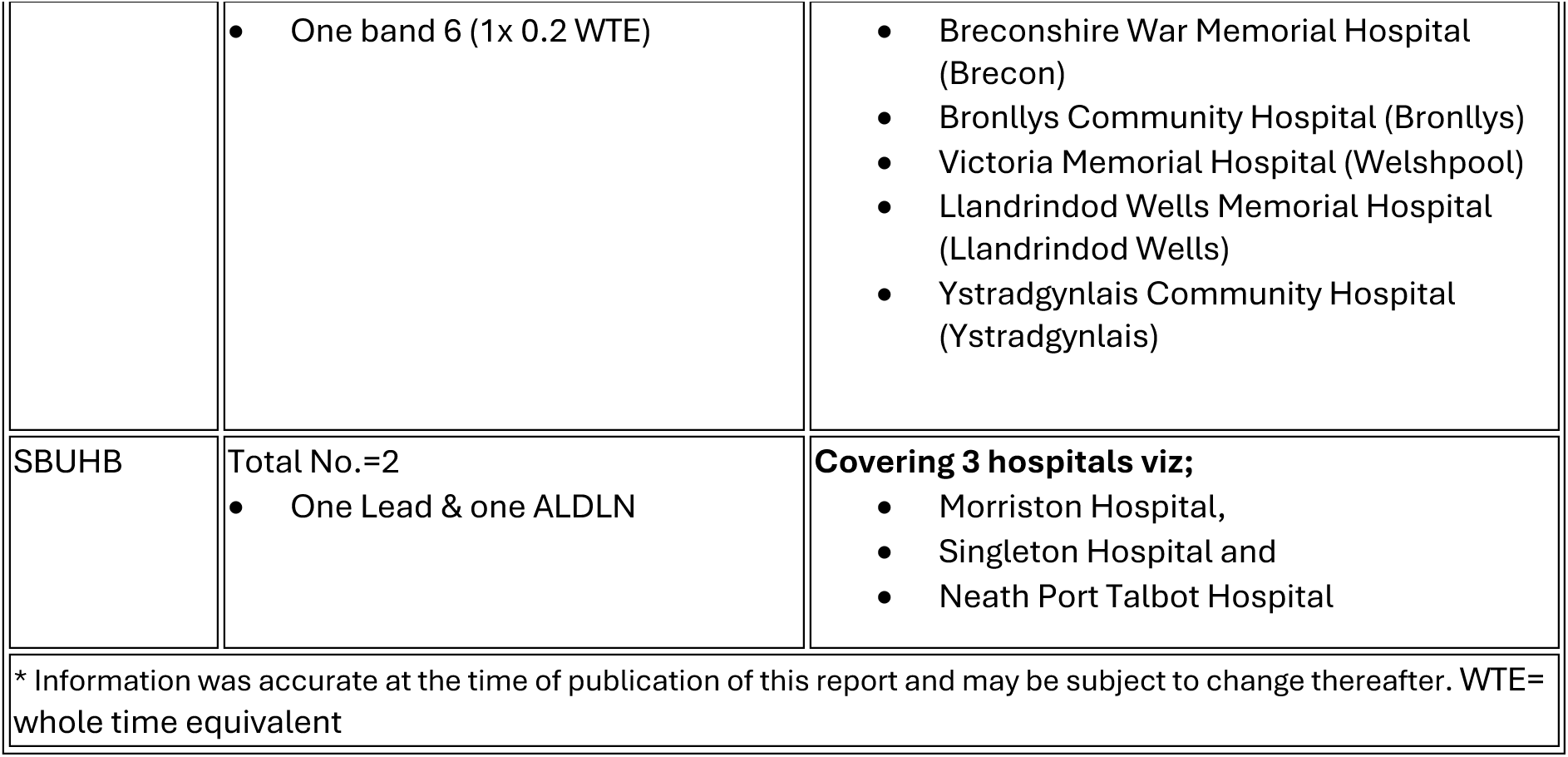
Current number of learning disability liaison nurses covering acute sites across the Welsh HBs*.

### 3.2 Overview of key themes

Framework analysis identified six main themes and thirty-five subthemes reflecting staff experiences of working with the ALDLS. These themes describe how the service operates in practice, the mechanisms through which it supports clinical teams and patients with a learning disability, and the organisational conditions that shape its impact across hospital settings. The themes are summarised in **Table *3*** and are presented below with illustrative participant quotations labelled by participants’ study ID and role.

**Table 3.**
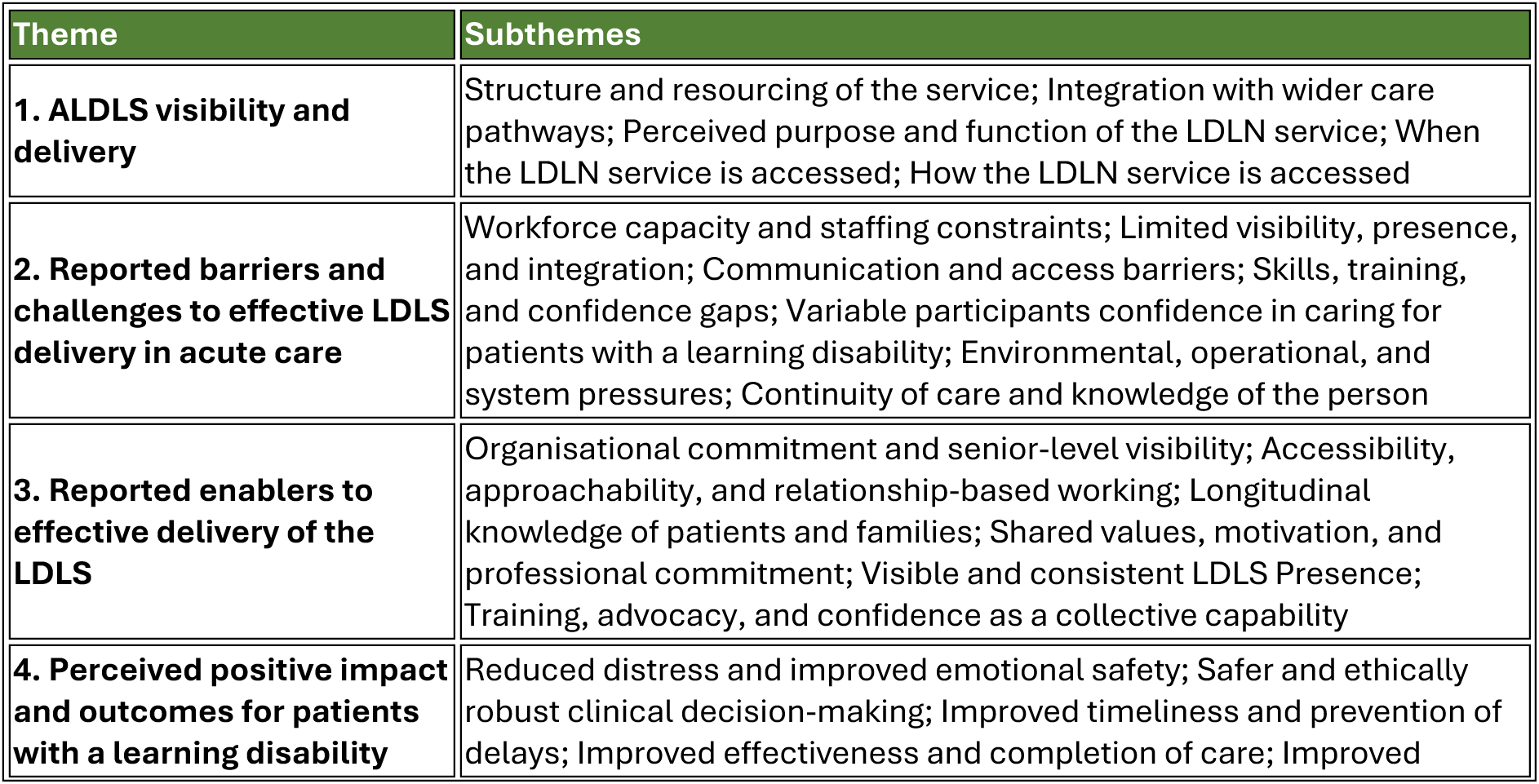
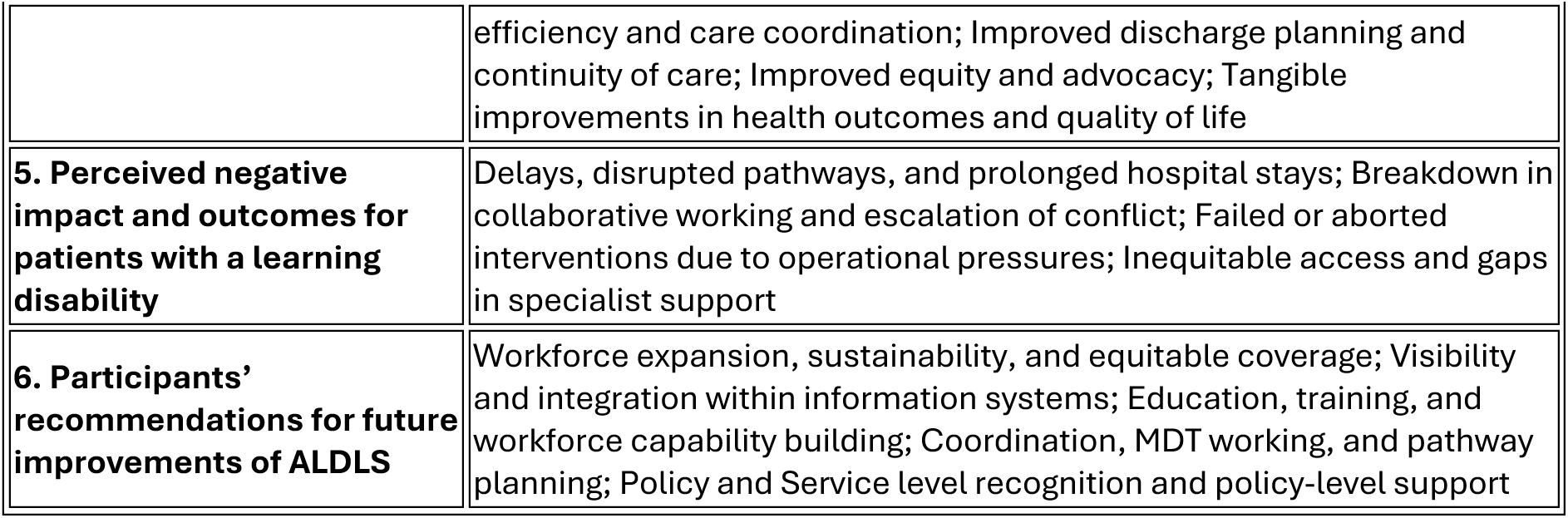
Overview of themes and subthemes.

| Theme | Subthemes |
| --- | --- |
| **1. ALDLS visibility and delivery** | Structure and resourcing of the service; Integration with wider care pathways; Perceived purpose and function of the LDLN service; When the LDLN service is accessed; How the LDLN service is accessed |
| **2. Reported barriers and challenges to effective LDLS delivery in acute care** | Workforce capacity and staffing constraints; Limited visibility, presence, and integration; Communication and access barriers; Skills, training, and confidence gaps; Variable participants confidence in caring for patients with a learning disability; Environmental, operational, and system pressures; Continuity of care and knowledge of the person |
| **3. Reported enablers to effective delivery of the LDLS** | Organisational commitment and senior-level visibility; Accessibility, approachability, and relationship-based working; Longitudinal knowledge of patients and families; Shared values, motivation, and professional commitment; Visible and consistent LDLS Presence; Training, advocacy, and confidence as a collective capability |
| **4. Perceived positive impact and outcomes for patients with a learning disability** | Reduced distress and improved emotional safety; Safer and ethically robust clinical decision-making; Improved timeliness and prevention of delays; Improved effectiveness and completion of care; Improved |
|  | efficiency and care coordination; Improved discharge planning and continuity of care; Improved equity and advocacy; Tangible improvements in health outcomes and quality of life |
| **5. Perceived negative impact and outcomes for patients with a learning disability** | Delays, disrupted pathways, and prolonged hospital stays; Breakdown in collaborative working and escalation of conflict; Failed or aborted interventions due to operational pressures; Inequitable access and gaps in specialist support |
| **6. Participants' recommendations for future improvements of ALDLS** | Workforce expansion, sustainability, and equitable coverage; Visibility and integration within information systems; Education, training, and workforce capability building; Coordination, MDT working, and pathway planning; Policy and Service level recognition and policy-level support |

### 3.3 ALDLS visibility and delivery

#### 3.3.1 Structure and resourcing of the service

Structural and resourcing constraints emerged as a central factor shaping perceptions of the service. Limited LDLS staffing, high caseloads, and coverage across multiple hospital sites restricted physical presence on wards and reduced opportunities for relationship-building with frontline staff. High turnover and loss of experienced staff were perceived to undermine continuity, institutional memory, and confidence in the service. As a result, the LDLN role was often experienced as peripheral rather than embedded within routine ward practice.

##### 3.3.1.1 Limited staffing and thin coverage

Participants consistently highlighted **under-resourcing**, with single nurses covering multiple hospitals, limiting visibility and consistency.

“There is now one nurse covering the three hospitals…she is too thinly spread.” *(P14_Sister Anaesthetics & Recovery and Part Time LD/Neurodiversity Lead Theatres)*

“There are not many of them, so their presence is limited.” *(P18_Senior Specialist Speech and Language Therapist)*

##### 3.3.1.2 Inconsistent physical presence on wards

The LDLN service was often described as **not routinely ward-based**, reducing opportunities for relationship-building and quick support to hospital staff.

“They rarely come on site; it is very rare to see them in person.” *(P26(FG)_Practice Development Sister_Emergency Department)*

“They are not routinely present on Wards, so you cannot just pop in and talk.” *(P18_Senior Specialist Speech and Language Therapist)*

##### 3.3.1.3 High turnover and loss of expertise

Several participants reflected on **loss of continuity and expertise** due to staff turnover and retirements.

“It is important to have sustainability; it is always nice to speak to the same people. If they know the patients really well over many years, that also really helps with continuity of care and building rapport with the patient.” *(P12_ Consultant in Special Care Dentistry)*

#### 3.3.2 Integration with wider care pathways

Integration with wider care pathways was described as variable. In some settings, LDLNs were well integrated into multidisciplinary teams, discharge planning, and elective pathways, effectively bridging community and acute care. Elsewhere, the service was perceived as external, with reliance on individual staff awareness rather than system-level identification mechanisms. While tools such as “The Once for Wales Health profile” (NHS Wales Performance and Improvement, ND) and electronic flags were recognised as valuable, their use was inconsistent, and staff education on their practical application was variable. Overall, the findings suggest that while the ALDLS is widely valued for its expertise and impact when engaged, its effectiveness is constrained by limited visibility, inconsistent access pathways, and structural pressures that limit full integration into acute care systems.

##### 3.3.2.1 Variable integration across health boards

Perceptions of integration varied widely, with some describing the service as embedded and others as external or standalone.

“These teams are treated as external to our work.” *(P25_Consultant in Acute Internal Medicine)* “Within the hospital that I am in, I think they are integrated as well as they can be.” *(P11_Nursing Associate (General Surgical Ward))*

##### 3.3.2.2 Bridging community and acute care

Where effective, LDLNs were valued for **bridging fragmented systems**, transferring knowledge from community to secondary care.

“This bridging work is vital.” *(P21_Heart Disease Specialist Nurse)*

“They have access to the community systems and can help us support patients.” *(P18_Senior Specialist Speech and Language Therapist)*

##### 3.3.2.3 Use of flags, passports, and profiles

Participants recognised hospital passports, flags, and The Once for Wales Health profile as **key tools**, though implementation was inconsistent.

“If a patient is known…a small banner appears on the clinical workstation.” *(P21_Heart Disease Specialist Nurse)*

“We used to have the butterfly symbol…it was much more visible.” *(P16_Pre-Op Assessment Clinics Nurse Practitioner)*

##### 3.3.2.4 Limited staff education and embedding

Several accounts suggested the ALDLS was **not fully embedded in workforce development**, limiting its system-wide impact.

“They were more visible on the ward…they would teach people how to fill out the forms.” *(P3_Clinical Nurse Specialist in Urology)*

“Because they’re not necessarily educating people…the impact is quite limited.” *(P3_Clinical Nurse Specialist in Urology)*

#### 3.3.3 Perceived purpose and function of the ALDLS

Across health boards, participants demonstrated a broadly shared understanding of the LDLN service as a specialist function focused on liaison, coordination, and the facilitation of reasonable adjustments for patients with a learning disability within acute care settings. The service was mostly described as a conduit between community learning disability services and hospital teams, supporting care planning, communication, and risk management, particularly for patients with complex needs or challenging behaviours. Where involved, LDLNs were valued for their expertise in understanding individual patients’ triggers, preferences, and support requirements, and for translating this knowledge into practical adjustments to care pathways.

##### 3.3.3.1 Liaison, coordination, and advocacy role

Participants commonly understood the ALDLS as a **liaison and coordination function**, acting as a bridge between acute services, specialist clinicians, and community LD teams, particularly for complex or high-impact patients.

“They shared with us the plans that the patient had set up in the community…and were able to share those resources with us.” *(P3_Clinical Nurse Specialist in Urology)*

“They meet people at the hospital door and escort them all the way through their appointments with us. Our patients are seen by healthcare professionals as well as pharmacists, and the liaison nurses accompany them through the whole appointment to support them as much as they can” *(P16_Pre-Op Assessment Clinics Nurse Practitioner)*

“To make sure patients feel listened to, cared for and valued, and that we are not speaking over them.” *(P23_Healthcare Support Worker)*

##### 3.3.3.2 Supporting reasonable adjustments and individualised care

A key perceived role was identifying and operationalising **reasonable adjustments**, including sensory needs, communication preferences, behaviour management, and least-restrictive care planning.

“She will go through a specific checklist to identify any reasonable adjustments for the patient’s hospital stay.” *(P12_Consultant in Special Care Dentistry)*

“They understand more than I do about communicating with a patient with learning disability.” *(P13_Band 7 Stroke Physiotherapist)*

##### 3.3.3.3 Practical rather than emotional support

Some participants framed the ALDLS as **practically focused**, supporting systems and processes rather than providing sustained emotional support to patients or staff.

“It was a less emotionally supportive service and more practically supportive service.” *(P3_Clinical Nurse Specialist in Urology)*

#### 3.3.4 When the ALDLS should be accessed

Understanding of *when* the service should be accessed was inconsistent. Many participants described LDLN involvement as largely reactive, often occurring after admission or in response to escalation, rather than being embedded early in the acute episode. Access was frequently described as threshold-based, with referrals more likely for patients presenting with behavioural challenges or requiring specialist interventions, rather than as a routine component of care for all patients with a learning disability. In contrast, planned pathways (such as elective procedures or multidisciplinary clinics) were more likely to benefit from proactive LDLN input, particularly where established relationships existed.

##### 3.3.4.1 Reactive and post-admission involvement

LDLN involvement was often described as **reactive**, occurring after admission rather than proactively during acute episodes, especially in emergency contexts.

“Engagement with LD tends to be after the event rather than during the acute clinical episode.” *(P26(FG)_Practice Development Sister_Emergency Department)*

“We only contact them if we need to; otherwise, we just pass each other in the corridor.” *(P23_Healthcare Support Worker)*

##### 3.3.4.2 Threshold-based or crisis-triggered access

Referral was frequently contingent on **severity, behavioural challenges, or clinical complexity**, rather than being routine for all patients with a learning disability.

“We only refer…if it’s behaviours which challenge, not at a basic level.” *(P1_Special School Nurse)*

“If it’s more complicated, then we need to bring in the PBM (Positive Behaviour Management) team.” *(P14_Sister Anaesthetics & Recovery and Part Time LD/Neurodiversity Lead Theatres)*

##### 3.3.4.3 Proactive involvement in planned pathways

In planned care (e.g. dentistry, anaesthetics), LDLNs were described as involved earlier, particularly through MDT working.

“Our MDT clinic is made up of the dental team, the anaesthetic team, and the learning disability nurse.” *(P12_Consultant in Special Care Dentistry)*

“We proactively involve them when planning to bring a patient into hospital for investigations.” *(P21_Heart Disease Specialist Nurse)*

#### 3.3.5 How the ALDLS is accessed

Participants also highlighted significant variability in *how* the service is accessed. Referral processes were often informal and ad hoc, relying on personal contacts, prior experience, or individual initiative rather than standardised pathways. Several staff reported uncertainty about referral criteria, responsibility for referral, and how patients were identified or flagged within hospital systems. While the service was widely described as responsive and accessible once contact was made, the lack of a clearly defined and consistently applied access mechanism limited its reach and reliability.

##### 3.3.5.1 Informal, ad hoc contact methods

Access was often described as **informal and inconsistent**, relying on personal contacts, emails, or previous interactions rather than a standardised system.

“I just e-mail a nurse…I find the last nurse I was talking to and I ask them, can I have help?” *(P2_Consultant Anaesthetist)*

“If somebody said to me…who do I contact, I wouldn’t know.” *(P8_Senior Nurse for Trauma)*

##### 3.3.5.2 Variable referral pathways and unclear responsibility

Participants expressed uncertainty about **who should refer, when, and through which system**, particularly across primary–secondary care boundaries.

“There is a lack of understanding about when to refer, who should refer.” *(P18_Senior Specialist Speech and Language Therapist)*

“I do not know how they know that patients are coming.” *(P16_Pre-Op Assessment Clinics Nurse Practitioner)*

##### 3.3.5.3 Accessibility once contact is made

Despite access challenges, once contacted the ALDLS was widely described as **responsive and helpful**.

“If it is urgent, I bleep her; she calls back and is normally on the ward within about 10 minutes.” *(P23_Healthcare Support Worker)*

“I’ve never been able not to be able to get support of the learning disability team.” *(P6_Ward Manager (Elective Admission Ward))*

### 3.4 Reported barriers and challenges to effective LDLS delivery in acute care

Participants identified a range of organisational and operational barriers that constrained the effective delivery and use of the ALDLS within acute hospital settings. Across sites, these barriers clustered around structural under-resourcing, limited-service visibility and integration within routine care pathways, and systemic gaps in staff training and confidence. Together, these factors had downstream effects on timeliness of care, patient safety, staff capability, and the overall experience of patients with a learning disability in hospital settings. While many participants recognised that these challenges reflected wider pressures within the health system, they were nevertheless viewed as potentially avoidable with clearer service models, improved integration, and strategic investment in workforce and training. A summary of the key barriers is presented in **Table 4**, followed by a detailed description of each subtheme with illustrative participant quotations in the section.

**Table 4.** Key barriers to effective delivery of the ALDLS.

| Barrier category | Description | Report Section | Page Number |
| --- | --- | --- | --- |
| **Workforce capacity and staffing constraints** | Limited LDLN staffing relative to demand restricts service coverage, responsiveness, and continuity, particularly across large hospital sites or outside core hours. | 3.4.1 | 30 |
| **Limited visibility, presence, and integration** | Participants reported variable awareness of the service and inconsistent integration within clinical teams, which reduced opportunities for early engagement and referral. | 3.4.2 | 30 |
| **Communication and access barriers** | Unclear referral pathways, reliance on informal contact, and inconsistent communication channels sometimes made the service difficult to access in practice. | 3.4.3 | 31 |
| **Skills, training, and staff confidence gaps** | Some staff reported limited training in caring for patients with a learning disability, resulting in uncertainty about appropriate adjustments and when to involve the LDLS. | 3.4.4 | 31 |
| **Variable participants confidence in caring for patients with LD** | Confidence in supporting patients with a learning disability varied across roles, departments, and levels of experience, influencing reliance on the liaison service. | 3.4.5 | 32 |
| **Environmental and operational pressures** | High patient throughput, time constraints, and competing clinical priorities limited the ability of staff to implement recommended adjustments or engage with the service. | 3.4.6 | 33 |
| **Continuity of care and knowledge of the person** | Fragmented care pathways and limited access to patient background information sometimes made it difficult to provide personalised support. | 3.4.7 | 33 |

#### 3.4.1 Workforce capacity and staffing constraints

The most frequently cited barrier was insufficient LDLN capacity relative to demand. Small teams covering large acute sites constrained responsiveness, visibility, and sustainability, often forcing prioritisation of only the most complex cases.

##### 3.4.1.1 Insufficient numbers of Learning Disability Liaison Nurses

Participants consistently described the LDLS workforce as too small to meet demand across large, multi-site acute hospitals. Limited staffing resulted in prioritisation of only the highest-risk cases, reduced visibility, and delayed engagement.

“This hospital is too big for two full time equivalent learning disability nurses…it is too big for two people because that’s what they’ve got.” *(P6_Ward Manager (Elective Admission Ward))*

“I think the barriers are not having enough learning disability nurses…they are particularly stretched and having to prioritise cases.” *(P12_Consultant in Special Care Dentistry)*

“I think there’s only one liaison nurse…the amount of liaison nurses that are available for the amount of learning disability patients we have is the challenge.” *(P9_Sister in Emergency Department)*

##### 3.4.1.2 Lack of cover during leave, sickness, and out-of-hours

The absence of systematic cover arrangements meant that LDLS support could become unavailable for extended periods, particularly affecting night shifts and weekends.

“If the nurse is off sick or on annual leave, there is not really anyone else to contact.” *(P17_Consultant in Special Care Dentistry)*

“Most of the time I work night shift…the service is only available Monday to Friday, 9:00 to 5:00.” *(P9_Sister In Emergency Department)*

“If she was off for two weeks, I don’t know whether anyone covers.” *(P14_Sister Anaesthetics & Recovery and Part Time LD/Neurodiversity Lead Theatres)*

#### 3.4.2 Limited visibility, presence, and integration within acute teams

##### 3.4.2.1 Perceived externality of the LDLS

Several participants described the LDLS as operating *alongside* rather than *within* acute services, contributing to delayed engagement and reduced ownership of shared care. “These teams are treated as external to our work…They do not feel integrated.” *(P25_Consultant in Acute Internal Medicine)*

“Engagement with LD tends to be after the event rather than during the acute clinical episode.” *(P26(FG)_Practice Development Sister_Emergency Department)*

##### 3.4.2.2 Reduced on-site presence and loss of routine touchpoints

Reduced ward attendance and absence from routine meetings limited relationship-building, awareness, and informal problem-solving.

*“They rarely come on site; it is very rare to see them in person.” (P26(FG)_Practice Development Sister_Emergency Department)*

*“We used to have representation there…but we don’t seem to have that anymore.” (P5_Senior Nurse (Acute Medical Unit))*

*“They only come probably once a week…if there were more of them, they could have more input.” (P13_Band 7 Stroke Physiotherapist)*

#### 3.4.3 Communication and access barriers

##### 3.4.3.1 Over-reliance on informal networks and personal relationships

While informal rapport facilitated access, participants highlighted this as fragile and unsustainable, especially during staff turnover.

“We had their personal phone number…that networking and rapport – although services should not rely on it – does matter.” *(P24 (FG)_Senior Nurse_Emergency Department)*

##### 3.4.3.2 Language barriers and lack of bilingual capacity

Language discordance, particularly for Welsh-speaking patients, was described as a significant barrier that compounded distress and reduced tolerance of care.

“Only one can speak Welsh…that is a problem.” *(P11_Nursing Associate (General Surgical Ward))*

“We had a dementia patient lately and they just could not tolerate speaking English.” *(P11_Nursing Associate (General Surgical Ward))*

#### 3.4.4 Skills, training, and confidence gaps among acute staff

##### 3.4.4.1 Limited staff confidence in implementing reasonable adjustments

Participants described uncertainty about what adjustments were permissible or effective, often linked to insufficient guidance and training.

“If someone said, what reasonable adjustment can you make…I wouldn’t know.” *(P3_Clinical Nurse Specialist in Urology)*

“What we can and can’t do, I wouldn’t know.” *(P3_Clinical Nurse Specialist in Urology)*

##### 3.4.4.2 Insufficient training on consent, capacity, and legal frameworks

Participants highlighted anxiety around consent, restraint, and litigation, leading to avoidance behaviours that compromised patient safety.

“I don’t feel that’s enough…their legalities, and consent.” *(P9_Sister In Emergency Department)*

“Staff avoid the patient…because they’re scared of litigation.” *(P9_Sister In Emergency Department)*

“You cannot say, ‘I’m going to restrain that patient’…we need specific guidelines.” *(P9_Sister In Emergency Department)*

“She was resisting it because she was very distressed, and the ward staff were trying to work out how to give her that treatment. They had called the dispensing team for advice, but in doing so the ward had not got it right and they were talking about holding her down to restrain her*.” ( P15_Mental Capacity Specialist Practitioner)*

#### 3.4.5 Variable participant confidence in caring for patients with a learning disability

##### 3.4.5.1 Confidence concentrated among senior staff and champions

Confidence in delivering independent care to patients with a learning disability without LDLN support was often highest among senior nurses, ward sisters, and designated LD champions, creating variability across teams.

“The ward sisters and Band-8 sisters are confident…maybe not all the healthcare assistants.” *(P23_Healthcare Support Worker)*

“Because I am a champion and have experience, I feel confident.” *(P19_Assistant Practitioner in a Surgical Assessment Ward)*

##### 3.4.5.2 Low confidence among general staff due to limited training

Participants highlighted widespread lack of confidence among general staff, particularly where formal training in LD care was limited or absent.

“I would probably say about 30% of staff would be confident…that’s why we need more training.” *(P9_Sister In Emergency Department)*

“We were not trained in this…having a learning disability team we can go to, helps us a lot.” *(P20_Unit Manager OPAMU (Older Persons Acute Medical Unit))*

##### 3.4.5.3 Staff can manage routine care but struggle with complexity

Participants reported that while routine care could often be delivered confidently, complex, unpredictable, or behavioural situations reduced confidence without LDLN involvement.

“I would feel somewhat confident…but I would not actually be able to do it.” *(P17_Consultant in Special Care Dentistry)*

“It is always a bit unknown; you never quite know how the appointment is going to go.” *(P16_Pre-Op Assessment Clinics Nurse Practitioner)*

##### 3.4.5.4 Confidence declines when LDLN presence or role clarity is reduced

Where LDLNs were perceived as less visible or less involved, staff reported lower confidence and greater reliance on their own experience.

“I do not feel particularly confident that the liaison team will support me…my confidence comes from my own experience.” *(P18_Senior Specialist Speech and Language Therapist)*

“I am still not entirely sure what they are aiming to do…it feels as though they are less present now.” *(P18_Senior Specialist Speech and Language Therapist)*

#### 3.4.6 Environmental, operational, and system pressures

##### 3.4.6.1 Acute care environments not conducive to LD needs

Busy wards, oversubscribed clinics, and time-pressured appointments limited the ability to provide individualised, patient-centred care.

“Our appointments are half an hour…it is not a long timeframe to do everything.” *(P16_Pre-Op Assessment Clinics Nurse Practitioner)*

“Hospitals are very busy…management cannot repeat that often because of service cost.” *(P17_Consultant in Special Care Dentistry)*

##### 3.4.6.2 Waiting lists and delayed access to treatment

Extended waits, particularly for theatre-based procedures, were described as unacceptable and disproportionately harmful for non-verbal patients.

“Some patients waited six years…many were in pain and non-verbal.” *(P17_Consultant in Special Care Dentistry)*

#### 3.4.7 Continuity of care and knowledge of the person

##### 3.4.7.1 Lack of consistent, person-centred knowledge

Frequent staff changes and limited time to build relationships reduced care quality and increased uncertainty.

“If staff keep changing and nobody really knows the patient, everything is up in the air.” *(P23_Healthcare Support Worker)*

“If you have one person who knows them ‘like the back of their hand,’ you will get somewhere.” *(P23_Healthcare Support Worker)*

### 3.5 Reported enablers to effective delivery of the ALDLS

Across sites, effective delivery of the Acute Learning Disability Liaison Service (ALDLS) was enabled less by formal structures alone and more by relational continuity, visibility, leadership endorsement, and values-driven practice. These enabling factors strengthened collaboration between liaison nurses and clinical teams and supported more confident care for patients with a learning disability. Where these enablers were present, staff confidence appeared more sustainable, particularly when managing complex or high-risk situations. A summary of the key enablers is presented in **Table 5**, followed by a detailed description of each subtheme with illustrative participant quotations.

**Table 5.** Key enablers supporting effective delivery of the ALDLS.

| Subthemes | Description | Report Section | Page Number |
| --- | --- | --- | --- |
| **Organisational commitment and senior-level visibility** | Leadership endorsement and policy and service level recognition of the service helped legitimise the role of LDLNs and supported integration of learning disability expertise within hospital systems. | 3.5.1 | 36 |
| **Accessibility and relationship-based working** | Participants valued the approachability and responsiveness of LDLNs, which fostered collaborative relationships and encouraged early engagement with the service. | 3.5.2 | 36 |
| **Longitudinal knowledge of patients and families** | Continuity of relationships enabled LDLNs to develop a deeper understanding of patients' needs, preferences, and communication styles, supporting more personalised care. | 3.5.3 | 37 |
| **Shared values and professional commitment** | A strong ethos of advocacy and person-centred care among LDLNs and wider staff supported collaborative problem-solving and improved patient experience. | 3.5.4 | 37 |
| **Visible and consistent LDLS Presence** | Regular presence on wards and consistent availability increased staff awareness of the service and facilitated timely access. | 3.5.5 | 38 |
| **Training, advocacy, and collective workforce capability** | Education, advice, and advocacy from LDLNs strengthened staff confidence and contributed to building a more capable workforce in caring for patients with a learning disability. | 3.5.6 | 39 |

#### 3.5.1 Organisational commitment and senior-level visibility

##### 3.5.1.1 Learning disability prioritised within organisational governance

Where learning disability was embedded into senior operational forums, staff described earlier identification of need, proactive planning, and clearer accountability for reasonable adjustments. “The moment an LD patient arrives we must make a plan…isolating them in a separate room…and contacting the LD nurses.” *(P24 (FG)_Senior Nurse_Emergency Department)*

“LD is firmly on our site-team radar so we can plan.” *(P24 (FG)_Senior Nurse_Emergency Department)*

##### 3.5.1.2 Inclusion of LD nurses in strategic and operational meetings

Early and routine involvement of LD nurses in meetings was seen as enabling smoother coordination and preventing reactive or fragmented care.

“She’s introduced into everything. It’s not as if she’s an afterthought…she’s always introduced right at the beginning of the meetings.” *(P4_Deputy Department Manager OPD)*

#### 3.5.2 Accessibility, approachability, and relationship-based working

##### 3.5.2.1 Approachability and ease of contact

Participants repeatedly identified approachability and responsiveness of LD nurses as central enablers of effective collaboration, supporting timely advice and confidence in decision-making. “They are always very approachable…very accessible, either by email or phone. That really helps.” *(P12_Consultant in Special Care Dentistry)*

“I see them as a crucial member of the team when planning for patients.” *(P12_Consultant in Special Care Dentistry)*

##### 3.5.2.2 Continuity of relationships over time

Sustained working relationships enabled deeper understanding of patients, smoother planning, and trust between teams.

“It is always nice to speak to the same people…If they know the patients really well over many years, that really helps with continuity of care.” *(P12_Consultant in Special Care Dentistry)*

#### 3.5.3 Longitudinal knowledge of patients and families

##### 3.5.3.1 Knowing patients beyond the acute episode

Participants described the value of long-term relationships with patients and families, allowing care to be tailored beyond immediate clinical needs.

“For us it is motivation and the nature of our relationships. We know our patients over a lifetime; they are not just ‘patient X’ but people we know, with families we know.” *(P21_Heart Disease Specialist Nurse)*

##### 3.5.3.2 Deep understanding of reasonable adjustments

Longstanding experience enabled staff to translate legislation and policy into practical, person-centred care.

“Over time…we have developed a strong understanding of what reasonable adjustments look like and how to connect guidance, protocols and legislation.” *(P21_Heart Disease Specialist Nurse)*

#### 3.5.4 Shared Values, Motivation, and Professional Commitment

##### 3.5.4.1 Values-driven practice and moral motivation

Strong professional values and prior exposure to poor care experiences motivated staff to champion improvement and innovation.

“Having witnessed poor examples of care…is a strong motivator to change and improve things.” *(P21_Heart Disease Specialist Nurse)*

“We care deeply and want to make a difference. Our knowledge helps us link with services and think about the bigger picture.” *(P21_Heart Disease Specialist Nurse)*

##### 3.5.4.2 Dynamic and proactive team culture

Teams described themselves as adaptive and opportunity-focused, actively seeking ways to improve care pathways.

“We are quite a dynamic team, always looking for opportunities.” *(P21_Heart Disease Specialist Nurse)*

#### 3.5.5 Visible and consistent LDLS presence

##### 3.5.5.1 On-site visibility as an enabler of engagement

Participants emphasised that visible and consistent presence (knowing when and where LD nurses would be available) enhanced utilisation and informal problem-solving.

“If you’re having one cover three main sites…do specific days…at least they could be visible on those days.” *(P8_Senior Nurse for Trauma)*

#### 3.5.6 Training, advocacy, and confidence as a collective, team-based capability

##### 3.5.6.1 Psychological safety from knowing specialist support is available

Across roles and settings, staff described increased confidence simply from knowing that LDLNs were available to advise, intervene, or provide reassurance. This “back-up” function reduced anxiety and supported decision-making, even among experienced clinicians.

“I’m confident because I know they’re there if I need them…now, yes, confident. But because I know they’re there.” *(P1_Special School Nurse)*

“All I have to do is pick up that phone…even if my brain is not working…they will support me in whatever I need.” *(P11_Nursing Associate (General Surgical Ward))*

“It’s just knowing they’re there at the end of the phone…that’s what we need.” *(P5_Senior Nurse (Acute Medical Unit))*

##### 3.5.6.2 Support for less confident, junior, or non-specialist staff

LDLNs were seen as particularly valuable for staff early in their careers or those without specialist LD training, helping to bridge confidence gaps.

“Dentists who have just come out of dental school…feel much less confident…the learning disability nurse is there to help colleagues who are less confident.” *(P12_Consultant in Special Care Dentistry)*

“For newer nurses, it would be a different story…there are still difficult situations that require intervention from the learning disability team.” *(P20_Unit Manager OPAMU (Older Persons Acute Medical Unit))*

##### 3.5.6.3 Confidence strengthened through team culture, shared learning, and reflection

Strong team cultures, psychological safety, and reflective practice enhanced confidence beyond individual competence.

“We share a strong value base…we do a lot of supervision and reflection.” *(P21_Heart Disease Specialist Nurse)*

“We are used to making reasonable adjustments…we try to teach new staff as they come along.” *(P6_Ward Manager (Elective Admission Ward))*

### 3.6 Perceived positive impact and outcomes for patients with a learning disability

Positive impacts were mapped analytically to the Institute of Medicine’s six domains of quality (STEEEP), with staff accounts illustrating improvements in safety, timeliness, effectiveness, efficiency, equity, and patient-centredness. Across settings, positive patient outcomes were not limited to clinical success, but included reduced distress, lawful care, improved access, smoother pathways, safer discharge, and better long-term quality of life. These outcomes were consistently linked to early LD liaison involvement, proactive planning, advocacy, and system-bridging work between services.

#### 3.6.1 Reduced distress and improved emotional safety during hospital care

##### 3.6.1.1 Tailored reasonable adjustments that minimise anxiety and escalation

Patients experienced reduced distress when health care environments, routines, and procedures were adapted to individual needs, often preventing behavioural escalation.

“If we say we need assistance with a particular procedure and how to do it with minimal distress, they are extremely helpful and give precise one-to-one advice.” *(P25(FG)_Consultant in Acute Internal Medicine_Inpatient)*

“We took every bit of orange out of his path…so there was not even a trigger before we even started.” *(P6_Ward Manager (Elective Admission Ward))*

“The smallest reasonable adjustment you can make, can make the biggest difference to that patient.” *(P6_Ward Manager (Elective Admission Ward))*

##### 3.6.1.2 Familiarity, routine, and desensitisation supporting patient tolerance

Gradual exposure, predictable routines, and personalised strategies enabled patients to tolerate hospital environments that would otherwise have been overwhelming.

“Over about six weeks…we gradually brought him nearer our department…using social stories and pictures.”

*(P14_Sister Anaesthetics & Recovery and Part Time LD/Neurodiversity Lead Theatres)* “Eventually he became more and more desensitised to the hospital. That was an achievement in itself.”

*(P14_Sister Anaesthetics & Recovery and Part Time LD/Neurodiversity Lead Theatres)*

#### 3.6.2 Safer, lawful, and ethically robust clinical decision-making

##### 3.6.2.1 Best-interest decision-making and lawful health care delivery

LD liaison input supported lawful, ethical health care, avoiding inappropriate restraint and ensuring decisions aligned with patients’ needs and rights.

“As a result, the person had the life-saving treatment she needed but was not restrained in a way that would have been completely inappropriate and unlawful.” *(P15_Mental Capacity Specialist Practitioner)*

*“They make sure we’re doing things properly and lawfully, not just what feels easiest at the time.”* (P12_Consultant in Special Care Dentistry)

##### 3.6.2.2 Structured risk assessment enabling appropriate care pathways

Structured risk assessments enabled patients to receive care safely while preserving dignity and autonomy.

*“If we had known that this patient tends to abscond, then we would have figured out other ways to make sure the patient was more comfortable and aware of what was going on.”* (P10_RN In Endoscopy)

“Without the learning disability team completing that risk assessment, I would have had conflict with the nursing staff.” *(P11_Nursing Associate (General Surgical Ward))*

#### 3.6.3 Improved timeliness and prevention of delays

##### 3.6.3.1 Enabling complex procedures through planning and coordination

LDLNs played a central role in enabling patients to access and complete procedures that might otherwise have failed.

“Our failure rates for general anaesthetic procedures are very low…due to good planning, good communication and strong MDT discussions.”

*(P17_Consultant in Special Care Dentistry)*

“That created the bridge between primary and secondary care…and was invaluable for safe admission and discharge.”

*(P17_Consultant in Special Care Dentistry)*

##### 3.6.3.2 Maximising clinical opportunities during admissions

Coordinated care ensured multiple health needs were addressed efficiently, reducing repeated admissions.

“We often do joint procedures…bloods, CT scans, ECGs…that has had a really positive outcome.”

*(P12_Consultant in Special Care Dentistry)*

LDLS engagement helped patients understand what to expect, reducing anxiety-related delays and improving cooperation with care processes.

*“If the patient understands what’s coming, they’re much more settled.”* (P10_RN In Endoscopy)

#### 3.6.4 Improved effectiveness and successful completion of care

##### 3.6.4.1 Improved communication for non-verbal or complex patients

LD teams supported interpretation of behaviours and symptoms, leading to more accurate diagnoses and care.

“We needed help to understand whether his non-verbal behaviours were pointing to dental pain.”

*(P17_Consultant in Special Care Dentistry)*

“When the learning disability team shared all their information with us, we gained a better idea of how to manage the patient.”

(P20_Unit Manager OPAMU (Older Persons Acute Medical Unit))

##### 3.6.4.2 Early identification and information sharing improving health care quality

Advance knowledge of patient needs allowed staff to prepare and deliver care more effectively. “Before the patient arrives we already know a lot, which makes our care easier and faster.” *(P19_Assistant Practitioner in a Surgical Assessment Ward)*

“They sometimes know about a patient before we do.”

*(P19_Assistant Practitioner in a Surgical Assessment Ward)*

#### 3.6.5 Improved efficiency and care coordination

##### 3.6.5.1 Communication and documentation

Centralised communication through the LDLS reduced repeated information requests and clarified care plans, allowing staff to focus on delivery rather than coordination.

*“It stops everyone asking the same questions over and over again.”* (P6_Ward Manager (Elective Admission Ward))

##### 3.6.5.2 Coordination across services

The LDLS played a key role in linking acute teams with community and social care services, improving flow across organisational boundaries.

*“They know who to contact straight away, which saves us days.”* (P11_Nursing Associate (General Surgical Ward))

#### 3.6.6 Improved discharge planning and continuity of care

##### 3.6.6.1 Safer, coordinated discharge back to familiar environments

LD liaison input supported safe discharge planning and prevented inappropriate placements.

“All of that planning was done by the learning disability team, which made it safer for the patient to go home.”

*(P20_Unit Manager OPAMU* (Older Persons Acute Medical Unit))

“Because plans were put in place, the patient could return to a familiar place.”

*(P20_Unit Manager OPAMU* (Older Persons Acute Medical Unit))

##### 3.6.6.2 Continuity across admissions and services

Ongoing oversight improved long-term care trajectories for patients with repeated admissions. “Having someone overseeing their journey was really helpful…ward nurses don’t have the time to follow up.”

*(P3_Clinical Nurse Specialist in Urology)*

“That extra layer meant the patient’s holistic needs were assessed.”

*(P3_Clinical Nurse Specialist in Urology)*

#### 3.6.7 Tangible improvements in health outcomes and quality of life

##### 3.6.7.1 Improved rehabilitation, mobility, and recovery

Specialist support enabled patients to engage with rehabilitation and recover more effectively. “They helped manage the patient so he could engage in rehabilitation and leave hospital.” *(P13_Band 7 Stroke Physiotherapist)*

##### 3.6.7.2 Long-term quality-of-life gains

Successful interventions had lasting benefits beyond the hospital admission. “He had a new lease of life…out painting, doing community activities.”

*(P14_Sister Anaesthetics & Recovery and Part Time LD/Neurodiversity Lead Theatres)*

### 3.7 Perceived negative impact and outcomes for patients with a learning disability in secondary care

Across accounts, negative patient outcomes were primarily system-driven rather than intent-driven, arising from service gaps, poor integration, inconsistent information, operational delays, and breakdowns in collaborative working. These factors contributed to prolonged distress, delayed or aborted care, inequitable access to specialist support, and strained decision-making processes, with direct implications for patient experience and safety.

#### 3.7.1 Delays, disrupted pathways, and prolonged hospital stays

##### 3.7.1.1 Prolonged admissions due to service access and coordination failures

Patients experienced extended stays in acute settings when specialist services were unavailable, poorly integrated, or slow to engage, leading to uncertainty and delayed decision-making.

“I recently had a patient in ED for over 72 hours because we could not get services to engage, mainly due to access and lack of robust job planning.” *(P24 (FG)_Senior Nurse_Emergency Department)*

“There were days of discussions about whether to scan, whether they needed a GA, and so on.”

*(P24 (FG)_Senior Nurse_Emergency Department)*

##### 3.7.1.2 Absence or reduced availability of LD liaison services resulting in patient suffering

Where LD liaison services were not readily available, participants perceived direct negative consequences for patients’ wellbeing.

“I think probably our patients are suffering at the moment because we haven’t got that service readily available or present.”

*(P8_Senior Nurse for Trauma)*

“I can imagine some patients are suffering right now because we haven’t got that.”

*(P8_Senior Nurse for Trauma)*

#### 3.7.2 Breakdown in collaborative working and escalation of conflict

##### 3.7.2.1 Variable quality of patient information undermining health care quality

Inconsistent or poor-quality written information limited staff’s ability to understand patients’ needs, resulting in inefficiencies and missed opportunities for appropriate care.

“The written information that comes with patients ranges wildly from invaluable to useless. Sometimes it completely explains what is going on and tells you exactly what to do; other times you may as well not read it.” *(P25(FG)_Consultant in Acute Internal Medicine_Inpatient)*

##### 3.7.2.2 Adversarial interactions undermining trust without altering parient outcomes

Contentious interactions between LD teams and clinical staff created conflict that diverted attention away from patient-centred decision-making, with negative emotional and procedural consequences.

*“Neurologists and physicians judged that he was nearing the end of life and that we should support him and his family rather than do further scans. The LD team pushed back, quoting disability legislation and suggesting we were discriminating because of his LD, which became very contentious for weeks. The medical examiner and coroner involved (because it was a death in care), both concluded nothing else could have been done. External scrutiny was fine and understandable, but it could have been done less confrontationally – for example, by asking if one of their clinicians could review the case as a second opinion, rather than emailing accusations of discrimination.” (P25(FG)_Consultant in Acute Internal Medicine_Inpatient)*

#### 3.7.3 Failed or aborted interventions due to operational pressures

##### 3.7.3.1 Delays and anxiety contributing to unsuccessful procedures

Operational delays and heightened anxiety among staff and carers negatively affected patients’ ability to tolerate procedures, resulting in aborted care.

“There was a delay in the patient coming into hospital and us giving him his pre-med…timing is very important.”

*(P12_Consultant in Special Care Dentistry)*

“You could tell the team with him were very anxious, and I think patients with learning disability pick up on other people’s anxieties.”

*(P12_Consultant in Special Care Dentistry)*

##### 3.7.3.2 Limits to implementing reasonable adjustments in busy hospital environments

Despite planning, systemic pressures constrained the extent to which reasonable adjustments could be fully implemented.

“Even though we plan as best as we can, there can still be delays in executing the plan.”

*(P12_Consultant in Special Care Dentistry)*

“The parents were saying, “No, they won’t allow you to do that.” But we need to do that. That is really important. So, I was trying to explain the importance in doing this procedure to them at that time. It takes a couple of hours. Well, in an emergency department, a couple hours is critical. Do you know what I mean?”

*(P9_Sister In Emergency Department)*

#### 3.7.4 Inequitable access and gaps in specialist support

Despite the benefits of LDLS, reliance on referrals and variable staff knowledge created inequities in access to the service.

*“If staff don’t know the service exists, that patient won’t get the same support.”* (P5_Senior Nurse (Acute Medical Unit))

### 3.8 Participants’ recommendations for future improvements of ALDLS

Across participants, future improvements centred on moving from a fragile, reactive, and peripheral model of LDLN provision to one that is adequately resourced, embedded, visible, digitally enabled, and strategically valued. Recommendations emphasised infrastructure, clarity, and capability-building, recognising that LDLNs cannot meet rising demand through individual effort alone.

#### 3.8.1 Workforce expansion, sustainability, and equitable coverage

##### 3.8.1.1 Increasing LDLN staffing levels to meet demand and reduce service fragility

Across health boards, participants consistently identified **insufficient staffing** as the primary constraint on LDLN effectiveness. Small teams covering large populations limited visibility, responsiveness, and continuity, creating a fragile service model vulnerable to sickness, vacancies, and burnout.

“There are only two nurses for all of UHW…they are excellent, motivated, knowledgeable and wonderful, but there are not enough of them.”

*(P21_Heart Disease Specialist Nurse)*

“One person carrying all that responsibility is too much.”

*(P17_Consultant in Special Care Dentistry)*

“Anything I have said about it not working as effectively as it could is directly caused by it being a very under-resourced service.”

*(P15_Mental Capacity Specialist Practitioner)*

##### 3.8.1.2 Embedding LDLNs at site or departmental level rather than pan–health board coverage

Participants advocated for **site-specific or specialty-linked LDLNs**, arguing that stable relationships and local knowledge improve access, trust, and effectiveness compared with centrally deployed teams.

“It would be nice to have somebody linked to each site…at least you’d have a name to go to rather than ‘the team’.” *(P7_Assistant Divisional Nurse for Medicine Division)*

“With better job planning we should, at minimum, have one LD liaison nurse based in each acute hospital.” *(P24 (FG)_Senior Nurse_Emergency Department)*

#### 3.8.2 Visibility, presence, and integration of information systems

##### 3.8.2.1 Increasing physical presence and in-reach within wards and departments

Greater **on-site visibility** was repeatedly described as transformational, improving patient flow, staff confidence, and quality of care. Infrequent or remote contact limited LDLN impact. *“Someone on site! Very simple…visible, present.”(P8_Senior Nurse for Trauma)*

*“When I had the LD liaison nurse on speed dial and she based herself in our department, we could see patients and send them home the same day.” (P24 (FG)_Senior Nurse_Emergency Department)*

##### 3.8.2.2 Moving from peripheral to embedded team membership

Participants described LDLNs as often feeling “external” to acute services. Embedding them within routine ward rhythms (e.g. daily or weekly check-ins) was seen as key to normalising their role.

“It would help to have them present, even once a week…a brief debrief on LD patients’ experiences.” *(P25(FG)_Consultant in Acute Internal Medicine_Inpatient)*

“They might come once a week, if that…more presence would help patient care definitely.”

*(P13_Band 7 Stroke Physiotherapist)*

##### 3.8.2.3 Improving electronic flagging and automated notification systems

Participants advocated for **system-level solutions** to identify patients with a learning disability early, reducing reliance on manual referrals.

*“If every morning an automatic email went to the LD team listing inpatients with LD flags, that could help them prioritise.” (P25(FG)_Consultant in Acute Internal Medicine_Inpatient)*

*“That would take away the referral process.” (P3_Clinical Nurse Specialist in Urology)*

##### 3.8.2.4 Making patient information clearer, shorter, and more usable

Participants highlighted the need for **concise, actionable prompts** rather than lengthy documents that are unrealistic to read in busy environments.

“Ward staff often do not have time to read lots of information.”

*(P18_Senior Specialist Speech and Language Therapist)*

“Shorter, clearer prompts about what each person needs would help.”

*(P18_Senior Specialist Speech and Language Therapist)*

#### 3.8.3 Education, training, and workforce capability building

##### 3.8.3.1 Expanding mandatory and face-to-face LD training for all staff

Participants emphasised **education as foundational**, particularly for new staff and non-specialist areas, to improve baseline competence and reduce over-reliance on LDLNs.

“Every member of staff should have more training, and that would help the service.”

*(P11_Nursing Associate (General Surgical Ward))*

“People need more face-to-face training…particularly around consent and legalities.”

*(P9_Sister In Emergency Department)*

##### 3.8.3.2 Strengthening LD champions and link nurse models

LD champions were viewed as effective but inconsistently supported. Revitalising networks and protected time for champions was seen as a pragmatic way to spread expertise.

“Training more champions would be good…it stems from education.”

*(P14_Sister Anaesthetics & Recovery and Part Time LD/Neurodiversity Lead Theatres)*

“We used to have a network of champions…I don’t know whether there’s time for that anymore.”

*(P14_Sister Anaesthetics & Recovery and Part Time LD/Neurodiversity Lead Theatres)*

#### 3.8.4 Coordination, multi-disciplinary team (MDT) working, and pathway planning

##### 3.8.4.1 LDLNs as coordinators of MDT meetings and care planning

Participants consistently described the LDLN role as ideally suited to **coordinating MDTs**, leveraging specialist knowledge and system-wide contacts to prevent delays.

“It would be really helpful if they oversaw the coordination of meetings.”

*(P18_Senior Specialist Speech and Language Therapist)*

“That would prevent delays in decision-making and discharge.”

*(P18_Senior Specialist Speech and Language Therapist)*

##### 3.8.4.2 Strengthening transition and cross-boundary working

Gaps between paediatric and adult services, and between community and acute care, were identified as priority areas for improvement.

“Transition from paediatric to adult services is hugely challenging.”

*(P21_Heart Disease Specialist Nurse)*

“There needs to be that link up a bit more between primary and secondary care.”

*(P7_Assistant Divisional Nurse for Medicine Division)*

##### 3.8.4.3 Clarifying the LDLN role and when to involve the service

Uncertainty about **what LDLNs do**, **when to refer**, and **what support they provide** led to underuse, delayed referrals, or inappropriate expectations.

“The lack of clarity about their role is a concern…it would help to clarify their aims and role so that we know how to use them.”

*(P18_Senior Specialist Speech and Language Therapist)*

“I do not know how to refer; it is not something we are very aware of.”

*(P18_Senior Specialist Speech and Language Therapist)*

##### 3.8.4.4 Introducing tiered or prioritised referral systems

Participants recommended **dual-level referral models**, distinguishing between notification and urgent clinical support, to improve responsiveness and reduce bottlenecks.

*“There needs to be explicit recognition that there are LD patients, and then there are LD patients who urgently need input.” (P25(FG)_Consultant in Acute Internal Medicine_Inpatient)*

*“Phone calls often hit answerphones and callbacks are slow or absent.” (P25(FG)_Consultant in Acute Internal Medicine_Inpatient)*

#### 3.8.5 Policy and service level recognition, valuation, and policy-level support

##### 3.8.5.1 Elevating the status of ALDLSs as essential, not optional

Participants framed LDLNs as **equivalent to other specialist nursing roles**, arguing that under-valuation undermines sustainability.

“You would not be without a diabetic nurse…so why would you not have somebody in the hospital for people with a learning disability?”

*(P14_Sister Anaesthetics & Recovery and Part Time LD/Neurodiversity Lead Theatres)*

“We need to place more value on this. It is an essential service.”

*(P14_Sister Anaesthetics & Recovery and Part Time LD/Neurodiversity Lead Theatres)*

##### 3.8.5.2 Aligning expansion with clear strategic objectives

While expansion was widely supported, participants emphasised the importance of **focused, goal-driven growth** rather than unfocused scaling.

“It’s not just an overall expansion; it’s an expansion to deliver specific strategic goals.”

*(P7_Assistant Divisional Nurse for Medicine Division)*

## 4 DISCUSSION

This evaluation explored hospital staff perceptions and experiences of the ALDLS, focusing on how the service is understood, accessed, and operationalised within acute hospital settings, and the extent to which it influences staff confidence in caring for patients with a learning disability, patient experience, and health care quality. The findings indicate that the service is widely valued, aligns strongly with national quality and equity agendas, and can deliver substantial benefits for patients with a learning disability. However, these benefits are inconsistently realised due to variation in service configuration, limited workforce capacity, and variable integration within acute care pathways. The relationships between key findings and strategic recommendations are summarised in a conceptual model illustrating how the ALDLS operates within acute hospital systems, highlighting the interaction between service mechanisms, organisational enablers and barriers, and resulting impacts on health care quality (Error! Reference source not found.**Figure 2**).

**Figure 2.**
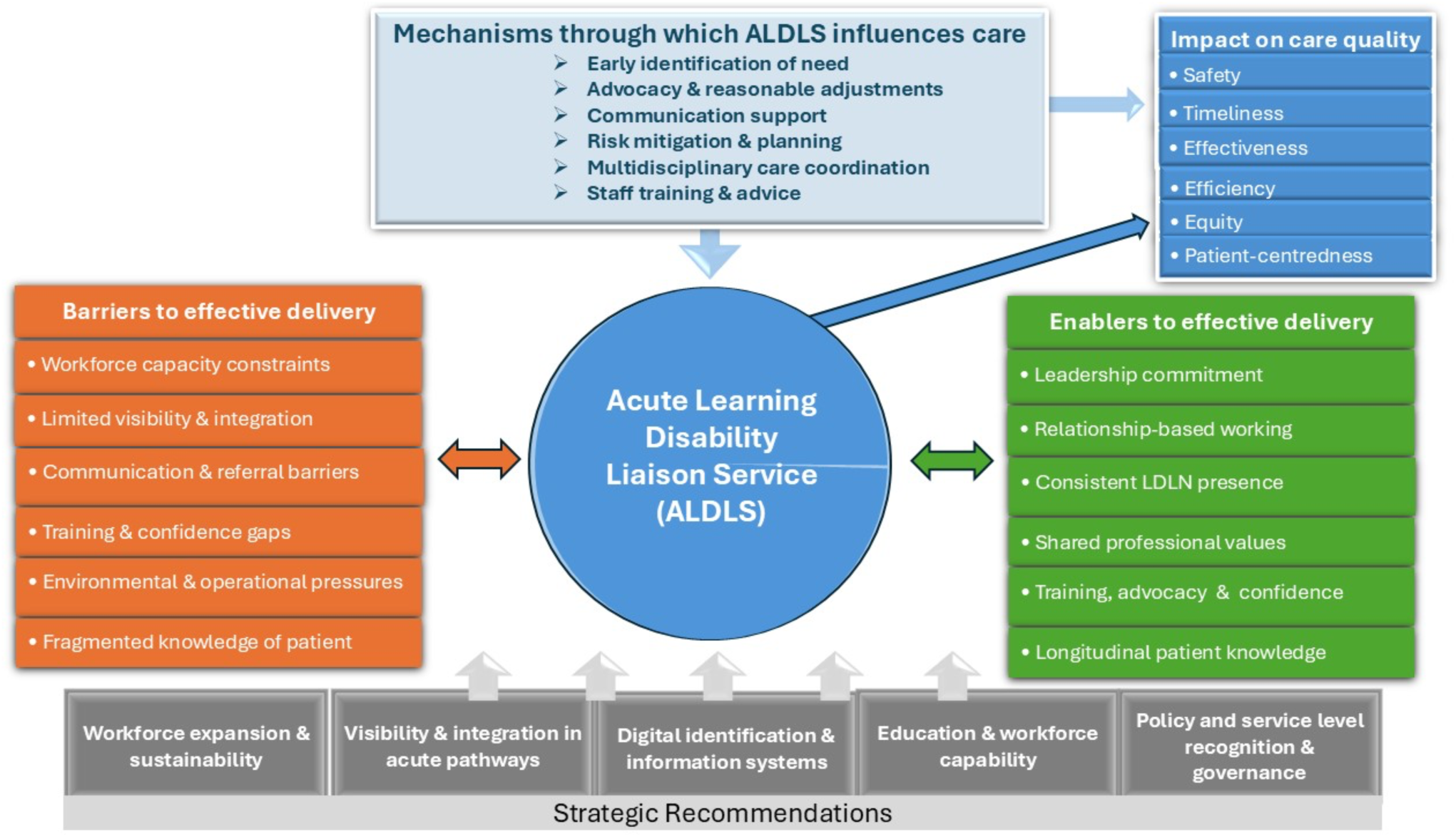
Conceptual model of how Acute Learning Disability Liaison Services influence care quality in acute hospitals.

In the discussion below, findings are synthesised across six main themes, highlighting key mechanisms through which LDLS adds value, as well as structural and systemic constraints that limit its reach and sustainability.

### 4.1 Understanding of the ALDLS and service configuration differences

#### Purpose, access, and configuration as determinants of impact

A central finding is that staff understanding of the LDLN role is highly context-dependent, shaped by how the service is configured and embedded locally. Where LDLNs were visible, routinely engaged in MDTs, and clearly positioned within acute pathways, staff articulated a coherent understanding of the service’s purpose—enabling reasonable adjustments, supporting decision-making, and advocating for patients. Conversely, where services were thinly spread or episodic in presence, understanding of the LDLN role was fragmented, leading to uncertainty about when and how to engage the service.

This finding is consistent with previous literature showing that liaison nursing roles are most effective when they are visible, accessible, and integrated into routine hospital practice rather than operating at the margins. Brown *et al*.(Brown et al., 2016) found that liaison nurses were valued for enabling person-centred care, advocacy, and reasonable adjustments, while Oulton *et al*. (Oulton et al., 2019) similarly reported that hospital staff perceived dedicated learning disability nurse provision positively, although provision was variable and unevenly embedded across services. Our findings extend this literature by showing that staff understanding of the service is not static or derived primarily from formal role descriptions; rather, it is actively co-produced through repeated contact, shared work, and local patterns of service presence. In this sense, variation in service configuration across Welsh Health Boards appears to shape not only access to the service, but also its perceived legitimacy and intelligibility within acute care.

#### Integration with wider care pathways

Limited integration between acute ALDLS provision and wider community, primary, and social care pathways further constrained effectiveness. Participants highlighted gaps at key transition points—particularly emergency admissions and discharge—where the absence of systematic information-sharing mechanisms placed additional burden on individual clinicians to ‘join up’ care. These findings reinforce the importance of the ALDLS not merely as a reactive, case-based service, but as a boundary-spanning function connecting traditionally siloed systems.

This resonates with wider evidence that people with a learning disability experience poorer care where reasonable adjustments depend on informal knowledge transfer rather than reliable systems, and where continuity across services is weak. Moloney *et.al* (Moloney et al., 2025) emphasise that implementation of reasonable adjustments in hospitals depends on mechanisms that support communication, coordination, and anticipatory planning, while Kelleher and colleagues (Kelleher et al., 2023) highlight the importance of joint working between acute and disability services in improving care for this group. Moloney *et.al* (Moloney et al., 2025) emphasised that implementation of reasonable adjustments in hospitals depends on mechanisms that support communication, coordination, and anticipatory planning, while Kelleher and colleagues (Kelleher et al., 2023) highlighted the importance of joint working between acute and disability services in improving care for this group. Our findings are broadly congruent with this literature, but add a more explicit organisational insight: hospital staff in Wales appeared to perceive the ALDLS as most effective when it functioned across boundaries (linking departments, services, and sectors) rather than as a stand-alone specialist resource. This suggests that the impact of liaison services depends not only on their specialist expertise, but also on the degree to which health systems enable them to act as connectors across fragmented pathways.

### 4.2 Perceived impact of the LDLS through the IOM six domains of quality framework lens

#### Safety, timeliness, and effectiveness as interlinked outcomes

Mapped against the Institute of Medicine’s six domains of quality framework, the LDLS was most consistently associated with improvements in **safety, timeliness, and person-centredness**, particularly through anticipatory planning and risk mitigation. These domains were not experienced independently; rather, timely LDLN involvement enabled safer care, which in turn supported more effective and less distressing clinical interventions.

This pattern is consistent with previous studies of learning disability liaison services, which have reported improvements in care coordination, communication, and implementation of reasonable adjustments when specialist liaison roles are available. For example, hospital staff in earlier research described liaison nurses as facilitating communication between patients, families, and clinical teams, and supporting safer and more coordinated care (Brown et al., 2016; Oulton et al., 2019). Similarly, qualitative research with liaison nurses has highlighted their role in advocating for reasonable adjustments, supporting clinical decision-making, and improving patient experience in acute care settings(Kelleher et al., 2025). Our findings reinforce these observations but add further insight by demonstrating how staff experience the domains of safety, timeliness, and effectiveness as closely interdependent rather than discrete outcomes. In practice, early LDLN involvement appeared to trigger a series of improvements including reducing uncertainty for staff, preventing escalation of distress in patients, and enabling clinical procedures to proceed more smoothly.

Notably, staff framed safety not only in terms of physical risk but also emotional and psychological safety, recognising distress, anxiety, and escalation as patient safety issues in their own right. This broader framing aligns with conceptualisations of patient safety that emphasise relational and contextual dimensions of care, particularly for patients with cognitive and communication differences (Bowness, 2014). It also resonates with evidence from UK (England) mortality reviews, which have repeatedly highlighted communication failures, diagnostic overshadowing, and unmet support needs as contributors to avoidable harm among people with a learning disability (Heslop et al., 2014; White et al., 2023)

#### Equity and efficiency as contingent benefits

Equity and efficiency gains were evident but more fragile, relying heavily on service availability and early identification of need. Where LDLNs were engaged proactively, staff described reduced duplication, smoother pathways, and fewer failed interventions. However, when access was delayed or inconsistent, inequities re-emerged, with patients experiencing prolonged stays, unmet needs, or repeated attempts at care.

These findings echo wider literature demonstrating that equitable hospital care for people with a learning disability depends heavily on the systematic implementation of reasonable adjustments rather than the goodwill or experience of individual clinicians(Moloney et al., 2023; Oulton et al., 2022). Studies examining the implementation of reasonable adjustments in hospital settings have similarly found that improvements in care quality often depend on reliable mechanisms for early identification and specialist support (Macarthur et al., 2015; Moloney et al., 2025). Our findings therefore reinforce the argument that equity gains are not simply the result of individual staff commitment but are contingent on organisational infrastructure—particularly the availability and integration of liaison services capable of coordinating adjustments and supporting staff decision-making. When such infrastructure is weak or inconsistent, existing inequalities in access, experience, and outcomes can quickly re-emerge.

### 4.3 Barriers and challenges to effective ldls delivery

#### Workforce constraints as the primary limiting factor

Across all sites and professional groups, **workforce capacity emerged as the dominant barrier** to effective LDLS delivery. Small teams covering large acute sites struggled to maintain visibility, provide timely input, and participate in routine ward activity. This often forced LDLNs into prioritising crisis response over upstream planning, thereby limiting their potential preventive impact.

Importantly, staff did not interpret these challenges as failings of individual practitioners, but as consequences of structural under-resourcing. This distinction is critical, as it underscores the risk of **over-reliance on professional goodwill** to compensate for systemic gaps. Similar workforce pressures have been reported in previous studies of learning disability liaison services, where limited staffing and competing demands constrained the ability of liaison nurses to engage proactively with wards and embed preventive approaches to care (Brown et al., 2012; Oulton et al., 2019). Our findings reinforce these concerns and suggest that insufficient staffing not only limits service reach but also shifts the focus of the role from anticipatory planning towards reactive crisis management, thereby reducing its potential impact on patient safety and care quality.

#### Visibility, communication, and environmental pressures

Limited physical presence within wards and departments compounded barriers created by workforce shortages. Many staff described uncertainty about who to contact, delays in responses, or reliance on generic inboxes, particularly in fast-paced environments such as emergency departments. These communication barriers were further exacerbated by acute operational pressures—bed shortages, staffing deficits, and time constraints—which reduced staff capacity to implement even well-designed reasonable adjustments.

These findings align with wider literature indicating that organisational pressures within acute hospitals can undermine the implementation of reasonable adjustments and coordinated care for patients with a learning disability (Moloney et al., 2025; Oulton et al., 2022). Previous research has similarly highlighted how high patient throughput, fragmented communication systems, and competing clinical priorities can limit staff abilities to respond effectively to the needs of this population (Heslop et al., 2013; White et al., 2022). In this context, liaison services are often expected to mitigate systemic constraints, yet their ability to do so depends heavily on their visibility and integration within routine clinical workflows.

#### Knowledge of the person as a fragile asset

A further barrier related to lack of continuity of care affecting knowledge of patients as individuals. High staff turnover, shift patterns, and fragmented documentation systems meant that personal knowledge (preferences, triggers, communication needs) was often lost or inconsistently applied. In this context, LDLNs played a critical role as carriers of longitudinal knowledge; however, their limited availability meant this function could not always be realised.

This observation is supported by previous research highlighting the importance of continuity of care in delivering person-centred care for people with a learning disability in hospital settings (Heslop et al., 2013). Studies show that when information about patients’ communication needs and support requirements is not systematically captured and shared across healthcare teams, care can become dependent on individual staff familiarity rather than reliable organisational processes (Heslop et al., 2013; Moloney et al., 2025). Our findings therefore reinforce the need for systems that support the consistent transfer of patient information (such as electronic flagging, The Once for Wales Health profile(NHS Wales Performance and Improvement, ND), and coordinated liaison support) so that personalised care does not rely solely on the presence of particular individuals.This observation is supported by previous research highlighting the importance of continuity of care in delivering person-centred care for people with a learning disability in hospital settings (Heslop et al., 2013). Studies have shown that when information about patients’ communication needs and support requirements is not systematically captured and shared across healthcare teams, care can become dependent on individual staff familiarity rather than reliable organisational processes (Heslop et al., 2013; Moloney et al., 2025). Our findings therefore reinforce the need for systems that support the consistent transfer of patient information (such as electronic flagging, The Once for Wales Health profile, and coordinated liaison support) so that personalised care does not rely solely on the presence of particular individuals.

Taken together, these barriers illustrate how the effectiveness of liaison services is shaped not only by professional expertise but by organisational infrastructure, workforce capacity, and information systems that enable (or constrain) the consistent delivery of reasonable adjustments in acute care.

### 4.4 Enablers to effective LDLS delivery

#### Organisational commitment and leadership visibility

Where senior leaders prioritised learning disability care (through mechanisms such as safety huddles, executive engagement, and formal escalation routes) LDLS involvement was normalised and legitimised. Organisational commitment acted as a sign of value, shaping staff behaviour and expectations around reasonable adjustments.

This finding is consistent with wider literature emphasising the role of organisational leadership and governance in improving hospital care for people with a learning disability. Previous research has highlighted that leadership commitment is critical to embedding reasonable adjustments and ensuring accountability for equitable care (Sheehan et al., 2016; Tuffrey-Wijne et al., 2014a). It was also found that liaison services are most effective when they are visibly supported by senior leadership and integrated into quality and safety governance structures, rather than operating as isolated specialist resources (Tuffrey-Wijne et al., 2014a; Tuffrey-Wijne et al., 2014b). Our findings reinforce the importance of leadership endorsement as a mechanism for legitimising the LDLS and signalling its relevance to everyday clinical practice.

#### Relationship-based working and shared values

Strong interpersonal relationships between LDLNs and acute staff consistently enabled effective care. Approachability, responsiveness, and continuity fostered trust and encouraged early engagement with the service. Shared professional values (particularly around dignity, equity, and advocacy) further reinforced collaboration, suggesting that culture and relational capital are as important as formal structures in enabling success.

These findings resonate with previous qualitative research demonstrating that the effectiveness of liaison roles often depends on relational working and informal networks within clinical teams (Kelleher et al., 2025). Liaison nurses have been described as “bridging” roles that rely on trust, accessibility, and collaborative problem-solving to influence practice across professional boundaries (Castles, 2012a; MacArthur et al., 2010). Our findings extend this work by highlighting how relational continuity not only facilitates communication but also supports the embedding of learning disability expertise within everyday ward culture.These findings are supported by previous qualitative research demonstrating that the effectiveness of liaison roles often depends on relational working and informal networks within clinical teams (Kelleher et al., 2025).

#### Predictable, consistent presence and training as catalysts

Predictable, regular LDLN presence (rather than ad hoc attendance) emerged as a critical enabler, allowing staff to integrate the service into routine care pathways. Training, advocacy, and peer encouragement also functioned as multipliers, extending LDLS influence beyond direct patient contact and embedding learning disability awareness more broadly across teams.

This observation aligns with evidence that education and specialist support can improve staff knowledge and confidence in caring for patients with a learning disability, particularly when training is reinforced through ongoing clinical collaboration (Tuffrey-Wijne et al., 2013). Previous studies have similarly emphasised that liaison nurses can act as educators and catalysts for organisational learning, helping to translate policy commitments around reasonable adjustments into practical changes in care delivery (Kelleher et al., 2025). Our findings suggest that a predictable, consistent service presence is particularly important in enabling this educational and cultural function.

#### Acute care staff confidence in caring for patients with a learning disability

Access to LDLNs significantly enhanced staff confidence, particularly in complex situations involving capacity, consent, distress, or behavioural escalation. However, confidence without specialist support was often described as fragile, contingent on individual experience and familiarity with the patient, or the presence of informal champions.

Confidence varied across staff roles, seniority, and clinical settings, with junior staff and those working in high-turnover environments reporting greater uncertainty. Importantly, participants conceptualised confidence not simply as an individual attribute but as a collective team capability, strengthened through shared learning, supervision, and psychological safety. This perspective aligns with wider literature indicating that staff confidence in caring for patients with a learning disability improves when specialist expertise, training opportunities, and supportive team cultures are present (Harrogate and District NHS Foundation Trust, 2017). Our findings therefore reinforce the idea that liaison services contribute not only to direct patient care but also to the development of a more capable and confident workforce.

### 4.5 Perceived impact and outcomes for patients with a learning disability

Across the dataset, positive impacts of the LDLS were described more often and in greater detail than negative ones, suggesting that when the service was accessible, visible, and involved early, it was widely perceived as making a meaningful difference to patient care and staff practice.

Participants gave rich examples of improved safety, reduced distress, successful completion of complex procedures, better discharge planning, and stronger advocacy for patients and families. These findings are broadly consistent with earlier research demonstrating that learning disability liaison services can improve communication, facilitate reasonable adjustments, and enhance patient experience within acute hospital settings (Hatton et al., 2011; Macarthur et al., 2015; Northway et al., 2024). Similarly, studies involving liaison nurses themselves have highlighted their role in advocating for patients, coordinating care, and supporting clinical teams to manage complex situations involving communication needs, consent, and behavioural distress (Bowness, 2014; Brown et al., 2012; Castles, 2012a).

However, negative experiences were also reported, and these were mainly linked to wider system pressures rather than isolated one-off events. When the service was short-staffed, unavailable out of hours, not well integrated into acute teams, or not clearly visible across sites, staff described delays, fragmented care pathways, avoidable distress, and unequal access to specialist support. These difficulties were most noticeable in large and busy hospitals with very small LDLS teams, in periods when posts were vacant, in evening and weekend care, and in areas where referral processes or role clarity were unclear. These findings mirror concerns raised in previous research, which have consistently identified gaps in coordination, communication, and access to reasonable adjustments as contributors to poorer hospital experiences and avoidable harm for people with a learning disability (Health Services Safety Investigations Body, 2023; Northway et al., 2024). Our findings reinforce this evidence by demonstrating how the presence (or absence) of liaison support can shape whether these systemic vulnerabilities are mitigated or amplified in everyday clinical practice.These findings mirror concerns raised in previous research, which have consistently identified gaps in coordination, communication, and access to reasonable adjustments as contributors to poorer hospital experiences and avoidable harm for people with a learning disability (Health Services Safety Investigations Body, 2023; Northway et al., 2024). Our findings reinforce this evidence by demonstrating how the presence (or absence) of liaison support can shape whether these systemic vulnerabilities are mitigated or amplified in everyday clinical practice.

### 4.6 Future improvements and strategic implications for LDLS services

#### From individual excellence to system reliability

Participants’ recommendations converge on a clear message: the LDLS must move from being a fragile, person-dependent service to a reliable, system-integrated function. Workforce expansion, equitable site coverage, and protected time for proactive planning are foundational requirements. These recommendations are consistent with previous research highlighting that liaison services often operate with limited staffing and high demand, constraining their ability to engage proactively with wards and participate in preventive planning (Health Services Safety Investigations Body, 2023; Moulster, 2020a). Our findings therefore reinforce the need to move beyond reliance on individual professional commitment and towards more stable service configurations capable of delivering consistent support across hospital settings.

#### Role clarity, training, and information systems

Clear articulation of LDLN roles, dual-tier referral pathways (notification vs urgent response), and improved electronic flagging systems were seen as essential to improving access and efficiency. Participants also highlighted mandatory training and strengthened learning disability champion networks as practical strategies for building baseline capability across the workforce while preserving specialist capacity for complex needs. These recommendations align with evidence demonstrating that effective implementation of reasonable adjustments requires systematic identification of patients with a learning disability, accessible patient information (Moulster, 2020a; Northway et al., 2024), and staff education supported by specialist expertise (MacArthur et al., 2010; Ní Riain & Wickham, 2024). Previous studies have similarly suggested that liaison nurses can act as catalysts for organisational learning by supporting training initiatives and promoting awareness of learning disability needs across clinical teams (Jennings, 2019; Tuffrey-Wijne et al., 2013). Strengthening these mechanisms may therefore help extend the influence of liaison services beyond individual patient encounters and contribute to wider positive cultural and organisational change.

#### Policy and service level recognition and policy alignment

Finally, the findings point to the need for explicit Policy and service level recognition of LDLS services within health board priorities and national policy frameworks. Participants consistently framed the service as essential to delivering equitable, safe, and person-centred care (objectives that align closely with national commitments to reduce health inequalities for people with a learning disability). Legislations such as the Equality Act 2010 (GOV.UK, 2010; Heslop et al., 2018), which requires healthcare providers to anticipate and implement reasonable adjustments for disabled people, and the ongoing learning from mortality review programmes (LeDeR) (White et al., 2026) both emphasise the importance of organisational systems that enable equitable access to care. Without sustained investment and policy-level support, the risk remains that LDLS impact will continue to depend on professional goodwill rather than system design. Strengthening and standardising LDLS provision may therefore represent a practical route to achieving equity of provision, improving patient safety, and addressing persistent disparities in healthcare outcomes for people with a learning disability.

### 4.7 Strengths and limitations

#### 4.7.1 Strengths

##### Comprehensive, multi-site qualitative dataset

A major strength of this evaluation is the breadth and depth of data collected across multiple acute hospital sites and all Welsh health boards. By including participants from a wide range of professional roles, specialties, and organisational contexts, the evaluation captures variation in how Learning Disability Liaison Services (LDLS) are understood, accessed, and enacted in practice. This enhances the **transferability** of findings across acute care settings with differing service configurations and levels of resourcing.

##### Rich, in-depth accounts of real-world practice

The use of semi-structured interviews and a focus group allowed participants to describe nuanced, experience-based perspectives on the operational realities of LDLS delivery. This approach enabled the evaluation to move beyond whether services exist to examine *how* they function, the mechanisms through which they add value, and the conditions under which they are most effective. The inclusion of illustrative quotations strengthens credibility by grounding analytic claims in participants’ own words.

##### Attention to both enabling and constraining factors

Unlike evaluations that focus solely on service effectiveness or deficits, this evaluation deliberately examined barriers, enablers and both positive and negative patient outcomes. This holistic approach allows for a more realistic appraisal of LDLS impact and avoids overly optimistic or problem-focused conclusions. The inclusion of future-oriented recommendations, grounded in frontline experience, further strengthens the utility of the evaluation.

##### Strong alignment with policy and practice priorities

The evaluation directly engages with national priorities around health inequalities, reasonable adjustments, patient safety, and workforce sustainability. By foregrounding issues such as workforce capacity, service integration, and equity of access, the findings are highly relevant to current NHS and Welsh Government policy agendas, enhancing the evaluation’s practical and strategic value.

#### 4.7.2 Limitations

##### Transferability but not generalisability from qualitative design

As a qualitative service evaluation, the findings are not intended to be statistically generalisable. While the inclusion of multiple sites and professional perspectives supports transferability, the results reflect perceptions and experiences rather than measured outcomes. However, this limitation is inherent to qualitative evaluation and is offset by the depth of insight gained into complex service processes.

##### Absence of direct patient and carer perspectives

This evaluation did not include primary data from people with a learning disability or their families and carers. This was a deliberate and proportionate design decision, informed by feasibility (including ethical approval process requirements and timelines) and the existing evidence base, which already robustly documents patient and carer experiences of hospital care and learning disability liaison services. Therefore, findings about positive or negative outcomes for patients must be viewed as perceived by staff, not actual evidence of outcomes for patients.

Given the rapid, time-limited nature of this evaluation, the focus was on hospital staff perspectives to examine how LDLS operates within acute systems and to identify practical service improvement opportunities. Meaningful involvement of patients and carers would have required additional time for accessible materials, consent processes, and support, and ethical approval processes, which were beyond the scope and required timelines of this work.

##### Variation in familiarity with LDLS among participants

Participants varied in their degree of exposure to and engagement with LDLS, which may have influenced their accounts. Some staff had long-standing relationships with LDLNs, while others had limited or episodic contact. Although this variation enriched the analysis by illuminating differences in service visibility and integration, it may also have shaped perceptions of service effectiveness and accessibility.

##### Potential for recall and attribution bias

As with all interview-based research, participants’ accounts may be influenced by recall bias or by particularly salient positive or negative experiences. Additionally, in complex care environments, it is sometimes difficult to disentangle the specific contribution of LDLS from other organisational or individual factors. The analytic framework sought to mitigate this by examining patterns across multiple accounts rather than relying on isolated examples.

##### Service evaluation context may limit critical disclosure

Because this work was conducted as a service evaluation rather than research, some participants may have been cautious in expressing criticism, particularly in relation to colleagues or organisational leadership. Nevertheless, the data included substantial critical reflection on service gaps, tensions, and negative outcomes, suggesting that participants felt sufficiently able to speak openly.

### 4.8 Implications for practice and policy

#### 4.8.1 Implications for practice

Findings highlight the ALDLS as a **core safety-critical function** within acute care, rather than an adjunct or optional service. Acute providers should therefore aim to embed ALDLS more visibly within routine clinical pathways, particularly in accident and emergency departments, day case surgery, and acute medical units where risks to people with a learning disability are greatest. Establishing predictable, consistent, site-based ALDLS presence (supported by clear referral criteria) would improve responsiveness, continuity, and staff confidence.

Practice should ideally move towards **proactive identification and early engagement** with ALDLS, supported by reliable electronic flagging systems and automatic notification processes. This would reduce reliance on individual staff awareness and prevent delays associated with reactive referrals. Structured MDT planning, including routine involvement of ALDLS in complex admissions and procedures requiring reasonable adjustments, should be standardised and systematised rather than dependent on local custom or individual advocacy.

The findings also demonstrate the need to **strengthen workforce capability** alongside specialist provision. Mandatory learning disability training (focused on consent, capacity, reasonable adjustments, and communication) should be complemented by ward-based champions and ongoing access to specialist advice. This approach would ensure that all or the majority of staff could potentially provide safe baseline care while recognising when specialist ALDLS input is required.

#### 4.8.2 Implications for policy

At policy level, ALDLS provision should be recognised as an **essential adjustment under equality and patient safety frameworks**, requiring sustained investment rather than short-term or discretionary funding. Variability in staffing levels, visibility, and coverage across sites undermines equity of access and contributes to preventable harm. Establishing national or regional minimum configuration standards for ALDLS (including baseline staffing levels, service coverage, core functions, and management and accountability arrangements) would help reduce this inconsistency and promote more equitable provision.

Workforce planning policies should reflect the **complexity and intensity of ALDLS work**, including the need for sufficient staffing to cover leave, sickness, and peak demand, and to allow time for governance, training, and system improvement activities. Consideration should also be given to extended or flexible service hours, particularly in emergency care settings.

Finally, national policy should support **integrated information systems** that enable consistent flagging of learning disability status and sharing of patients’ health profiles across care settings. Strengthening digital connection of clinical record systems between primary, community, and secondary care would improve continuity, reduce duplication, and support person-centred decision-making throughout the patient journey.

### 4.9 Recommendations

To translate the findings of this evaluation into practical change, the actionable recommendations outlined below (**Table 6**) synthesise staff-identified priorities into system-level interventions. These recommendations focus on strengthening workforce sustainability, improving integration into acute pathways, enhancing digital identification and coordination, and embedding learning disability expertise within governance and training structures. Together, they provide a structured roadmap for aligning ALDLS in secondary care with equality duties, patient safety standards, and quality improvement principles. Investment in these areas supports compliance with equality legislation, aligns with LeDeR safety learning, and advances all six IOM domains of health care quality.

**Table 6.**
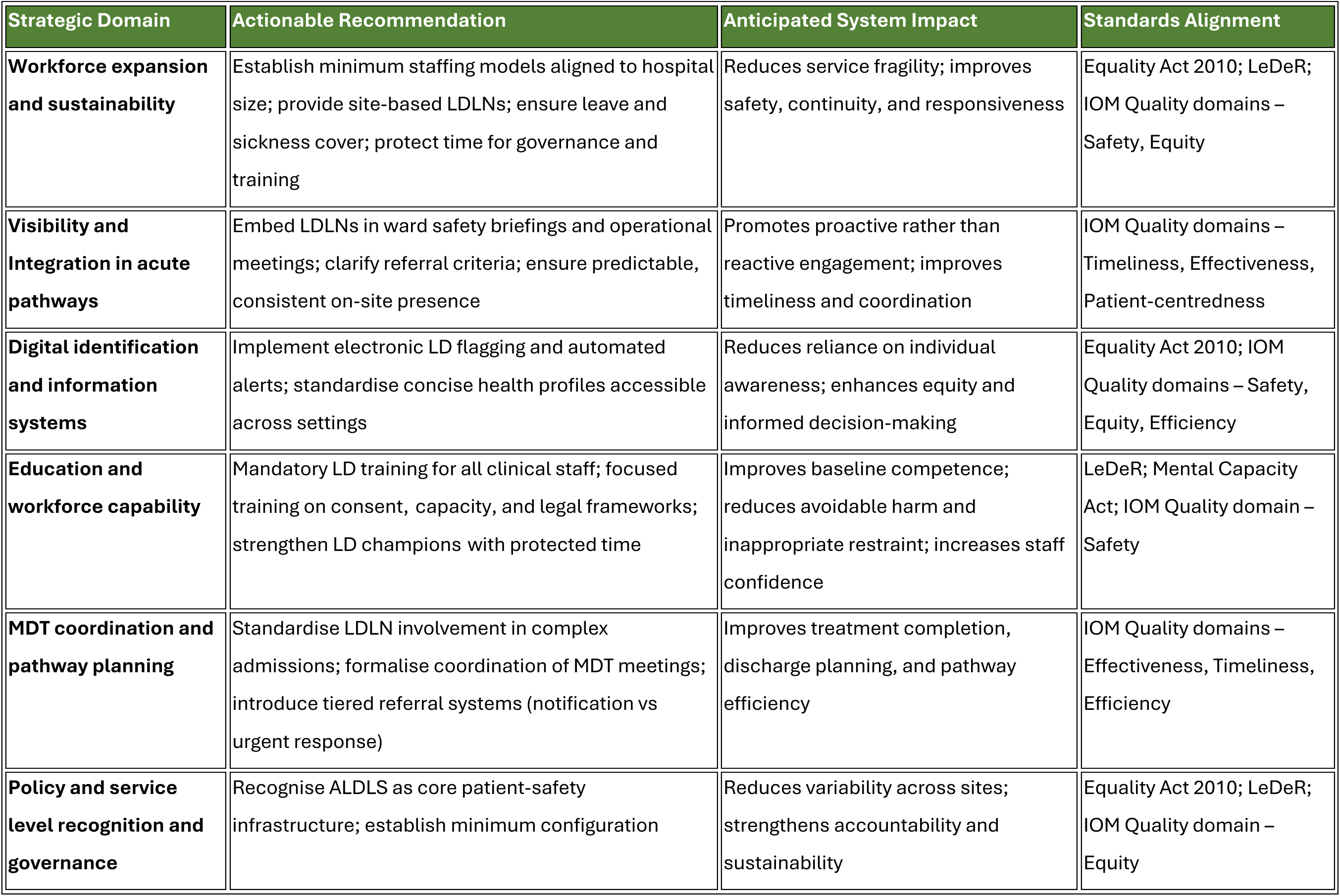
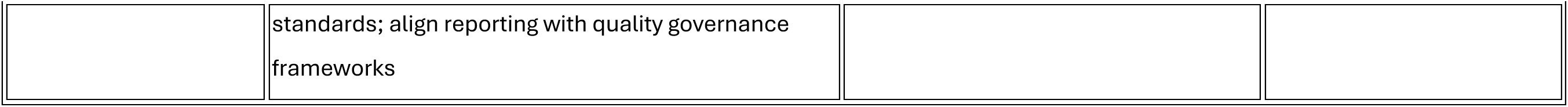
Summary of Practice and Policy Implications for Strengthening ALDLS in Acute Care.

### 4.10 Conclusions

This evaluation demonstrates that the ALDLS plays a critical role in enabling safe, equitable, and person-centred care for patients with a learning disability in acute hospital settings. Across health boards, hospital staff consistently described the ALDLS as a key mechanism for translating legal duties, professional values, and policy commitments into operational practice, particularly through early identification, advocacy, tailored reasonable adjustments, and coordination across complex care pathways.

Findings show that when the ALDLS is visible, integrated, and adequately resourced, it is reported to contribute towards improved hospital staff confidence and perceived to achieve more timely and effective clinical decision-making, reduced patient distress, safer admissions, and more coherent discharge planning. Conversely, variability in service configuration, limited workforce capacity, and inconsistent integration within acute teams were considered to undermine these benefits, exposing patients to avoidable delays, fragmented care, and inequitable experiences. Such gaps may also hinder the consistent implementation of the reasonable adjustments duty set out in the Equality Act 2010, which requires healthcare organisations to anticipate and remove barriers to equitable care for disabled people, including those with a learning disability.

Overall, the study highlights that ALDLS should not be viewed as optional or supplementary, but as a core component of high-quality acute care for people with a learning disability. Strengthening and standardising ALDLS provision (through sustainable resourcing, clear role definition, improved visibility to service users, and proactive integration into acute setting) offers a practical route to addressing persistent inequalities identified in this evaluation, and to delivering care that aligns with statutory obligations and established quality frameworks.

## Data Availability

All data produced in the present study are available upon reasonable request to the authors

## 6. ACKNOWLEDGEMENTS

The authors would like to thank the hospital staff who participated in the interviews and focus group, and who generously shared their experiences and perspectives. We are also sincerely grateful to the Acute Learning Disability Liaison Nurse Teams across all Welsh Health Boards for their time, involvement, and invaluable support with participant recruitment. In particular, we would like to acknowledge: Amy Bold, Andy Jones, Catherine Davies, Clare-Louise Owen, David Martin Lloyd, Emily Howells, Joanna Moyle, Melissa Evans, Neil James, Sharon Dixon, Simon Meadowcroft, Sonia Winley, Sophie Crabb, Wendy James. Their contributions were essential to informing this evaluation and enhancing understanding of the role and impact of Acute Learning Disability Liaison Services within hospital settings.

## 7. APPENDIX

## 8 LIST OF APPENDICES

|  |  |
| --- | --- |
| Appendix 1 | Interview recruitment poster |
| Appendix 2 | Participant information sheet |
| Appendix 3 | Screening and consent |
| Appendix 4 | Focus group recruitment poster |
| Appendix 5 | Interview guide |
| Appendix 6 | Focus group facilitator guide |

## 8.1 Appendix 1. Interview recruitment poster

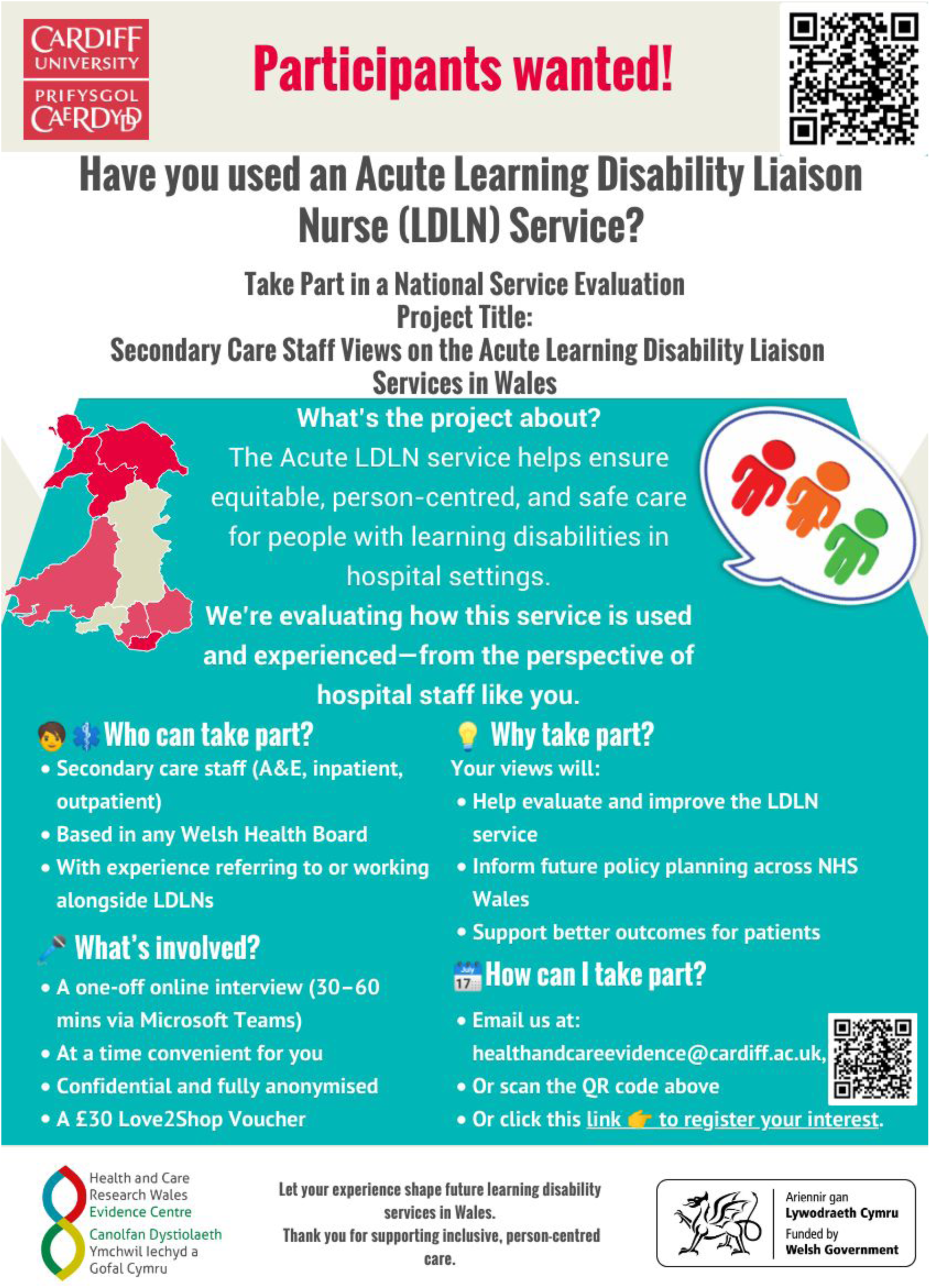

## 8.2 Appendix 2. Participant information sheet

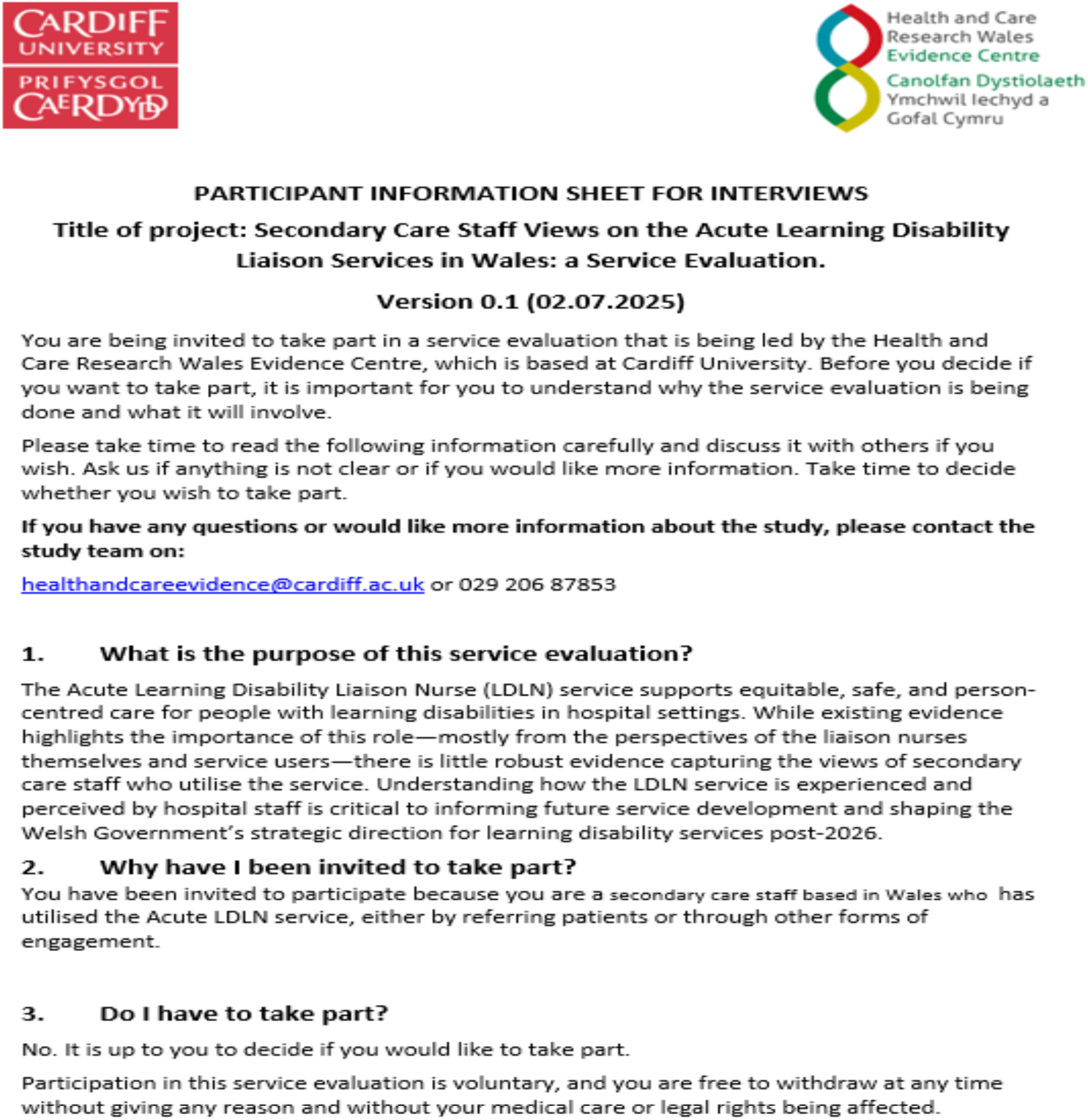
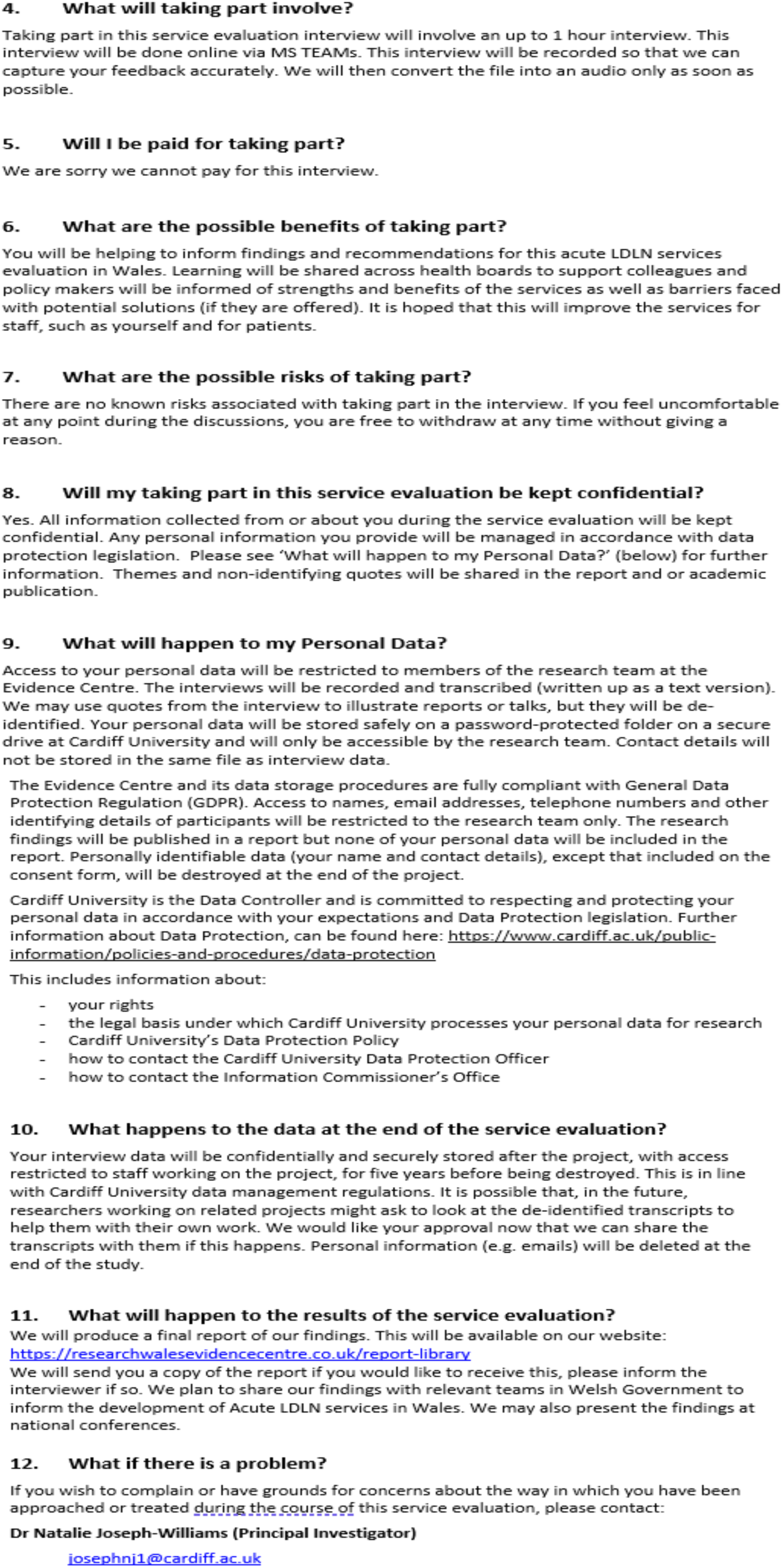
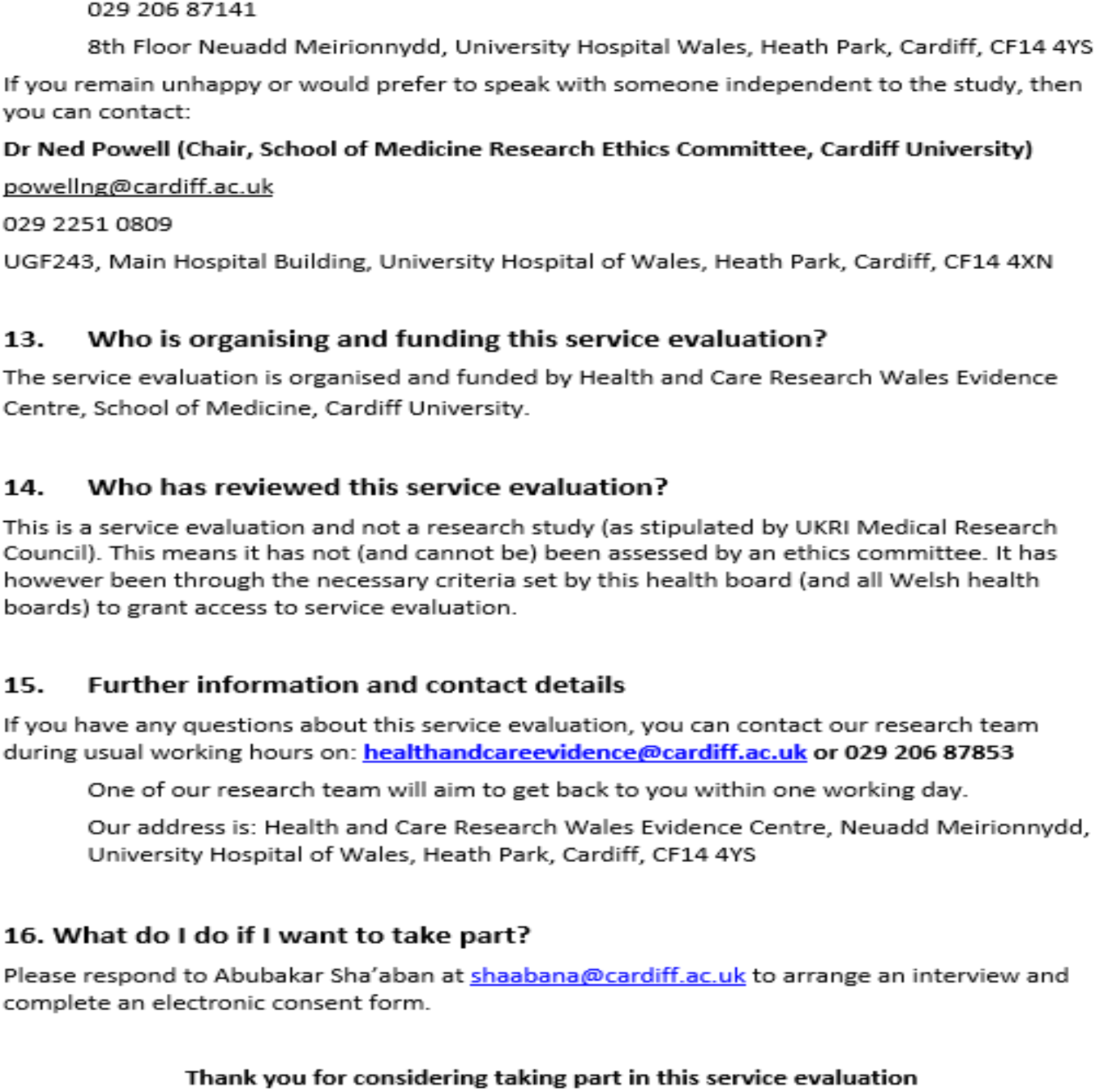

## 8.3 Appendix 3. Screening and consent form

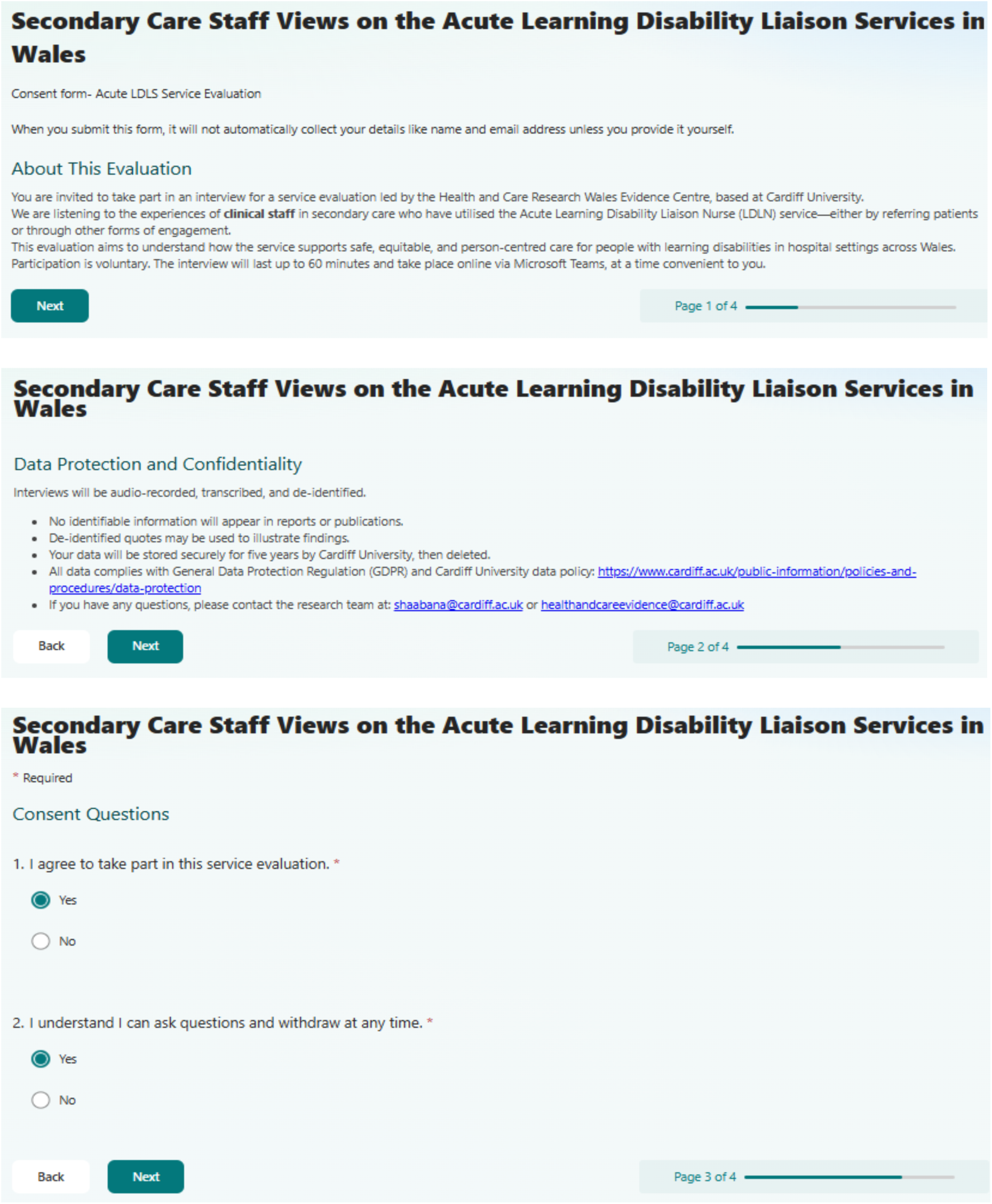
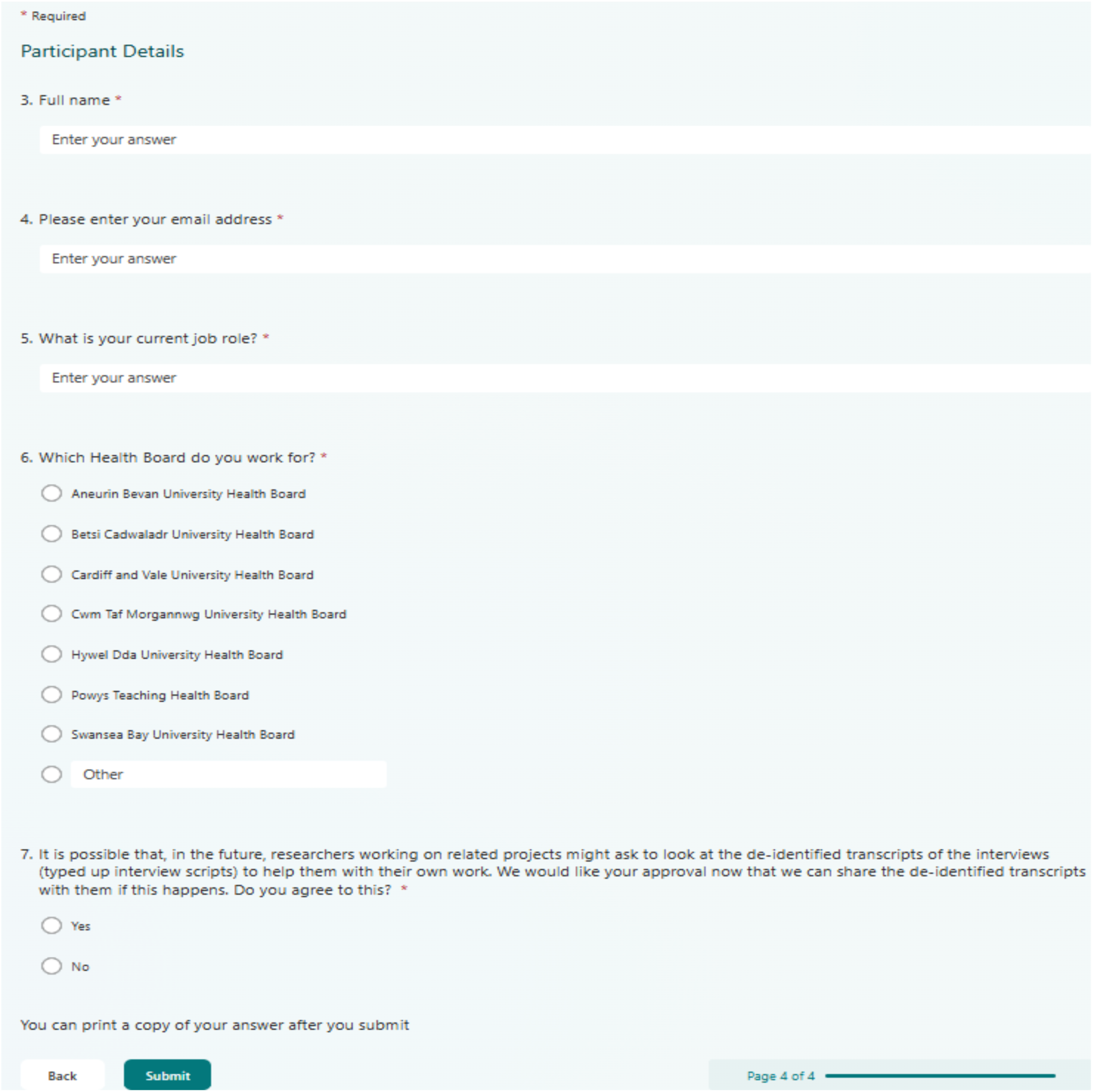

## 8.4 Appendix 4. Focus group recruitment poster

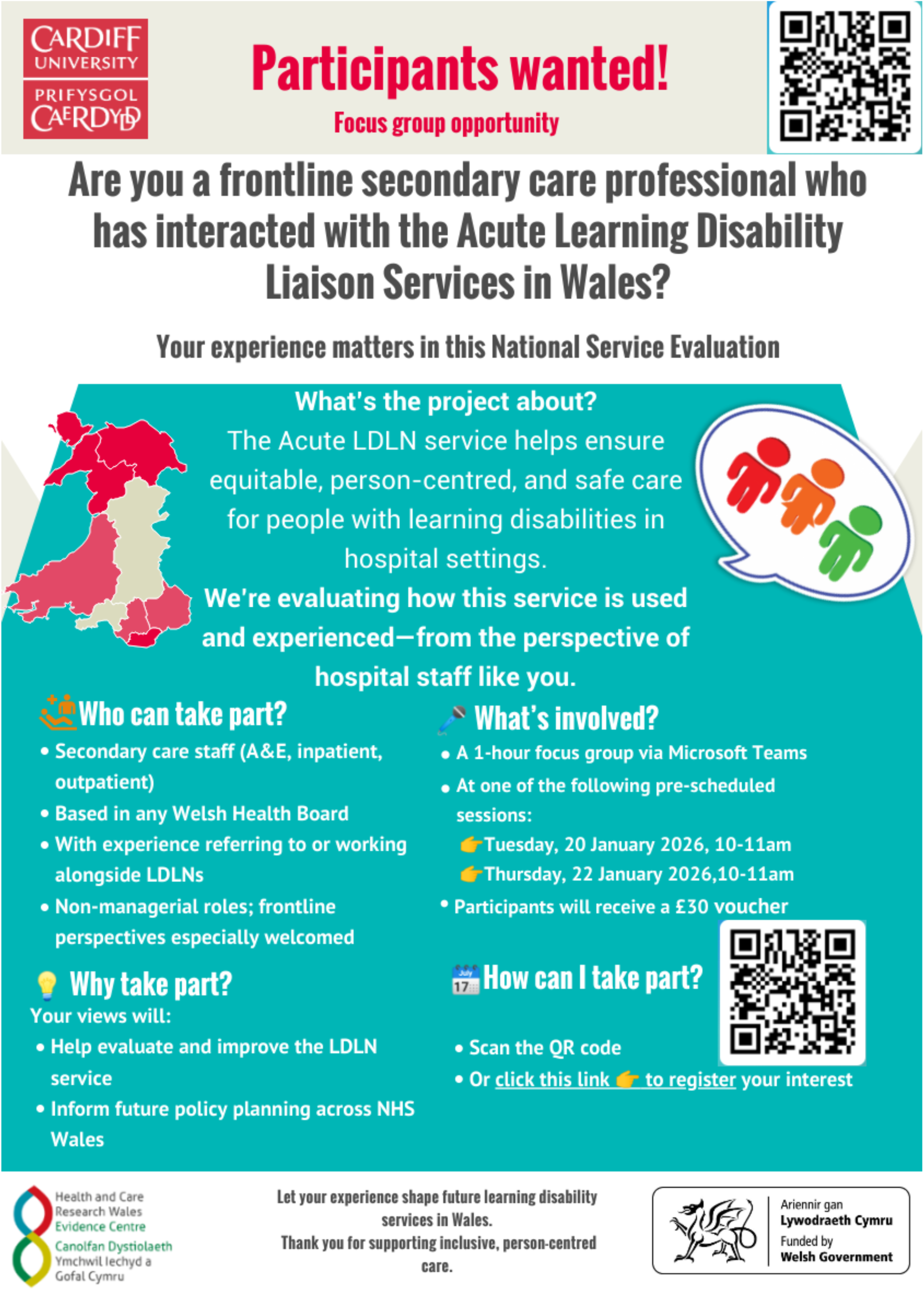

## 8.5 Appendix 5. Interview guide

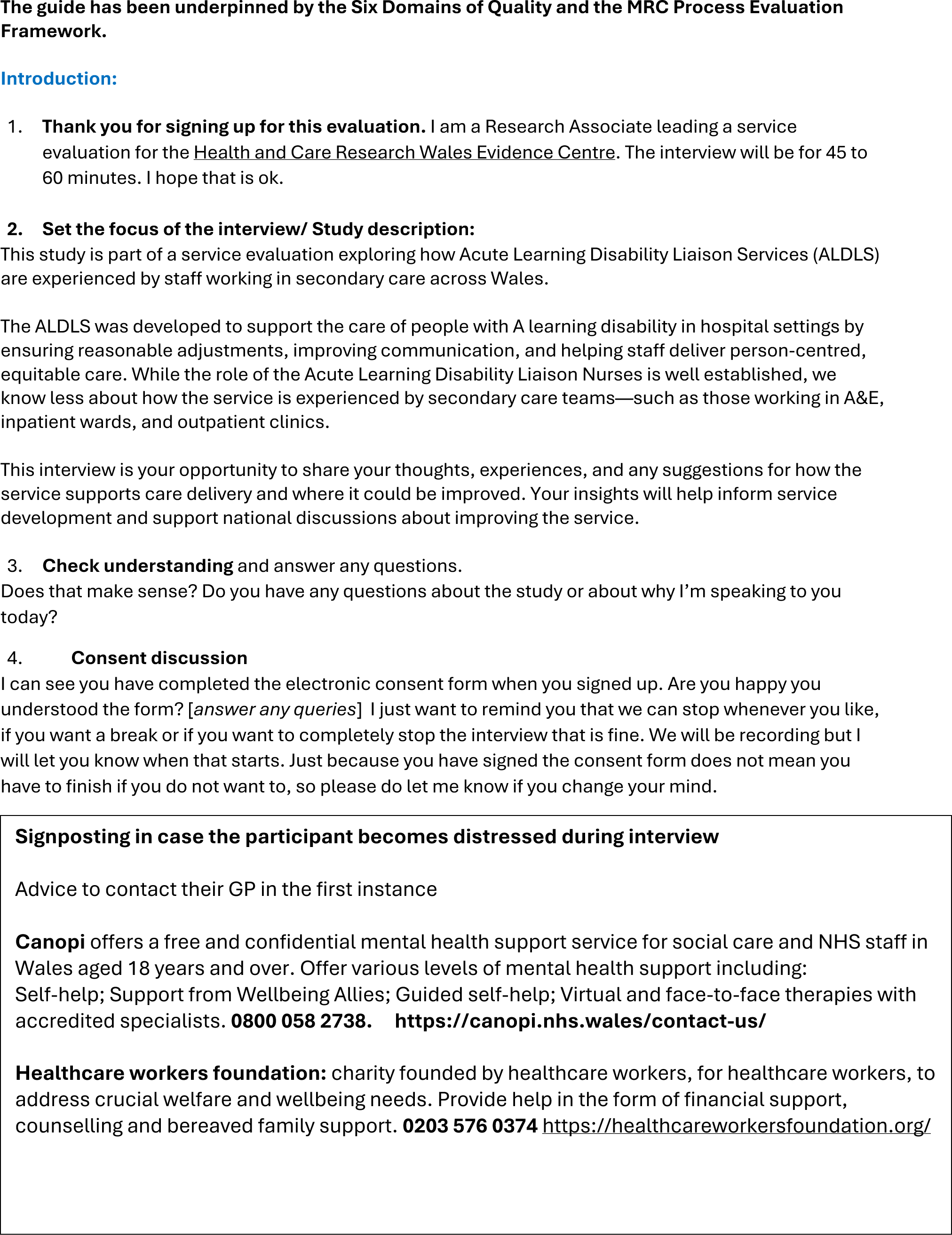
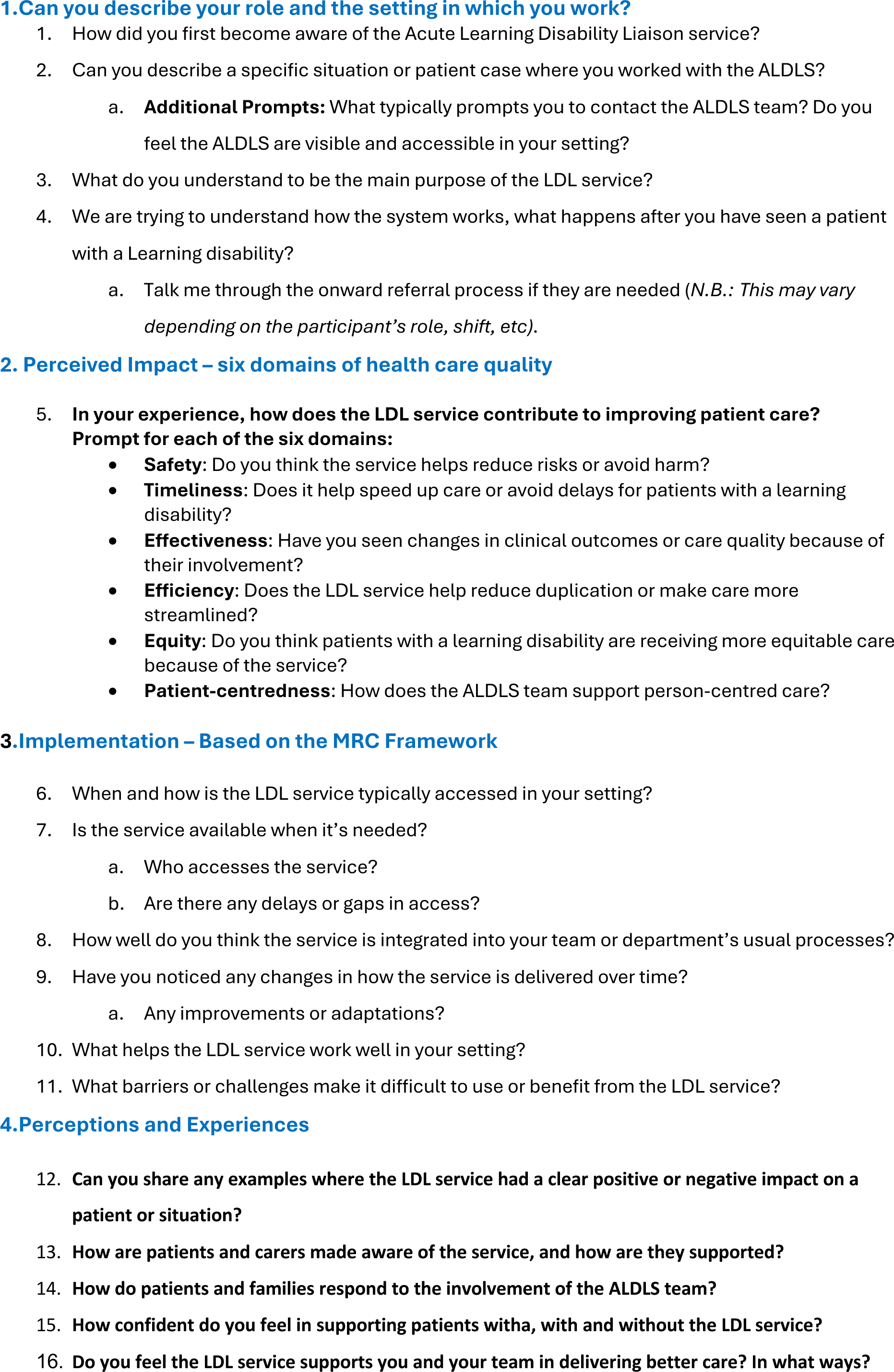
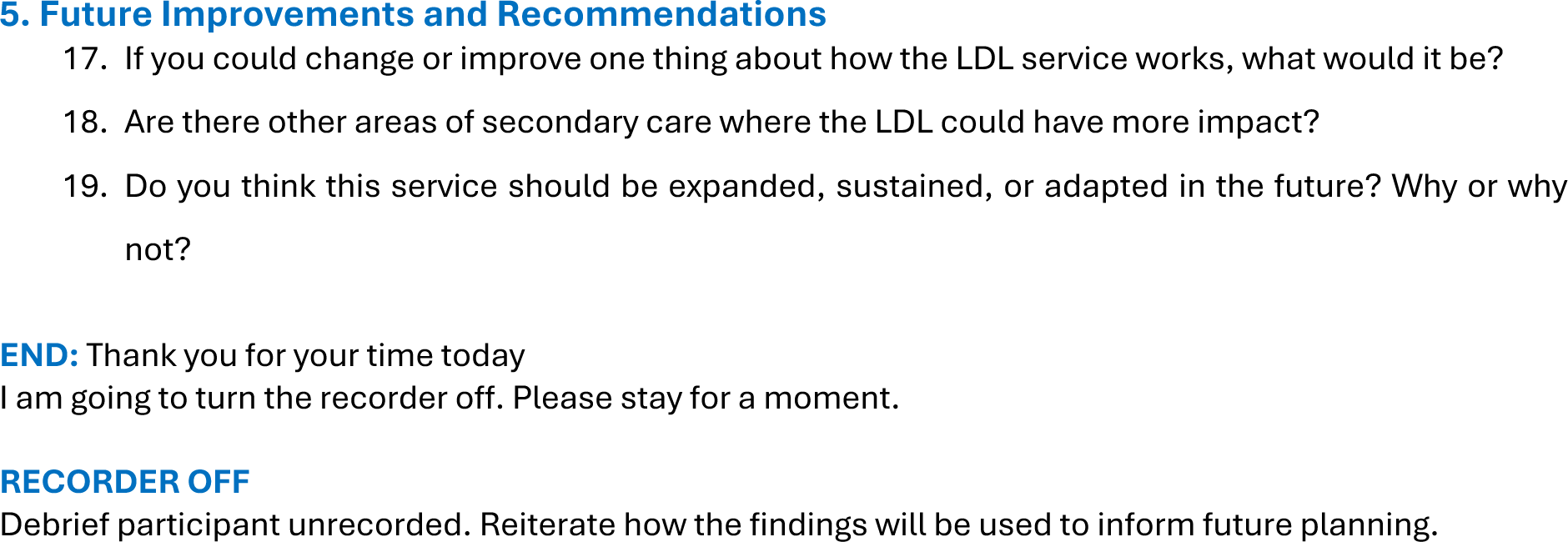

## 8.6 Appendix 6. Focus group facilitator guide

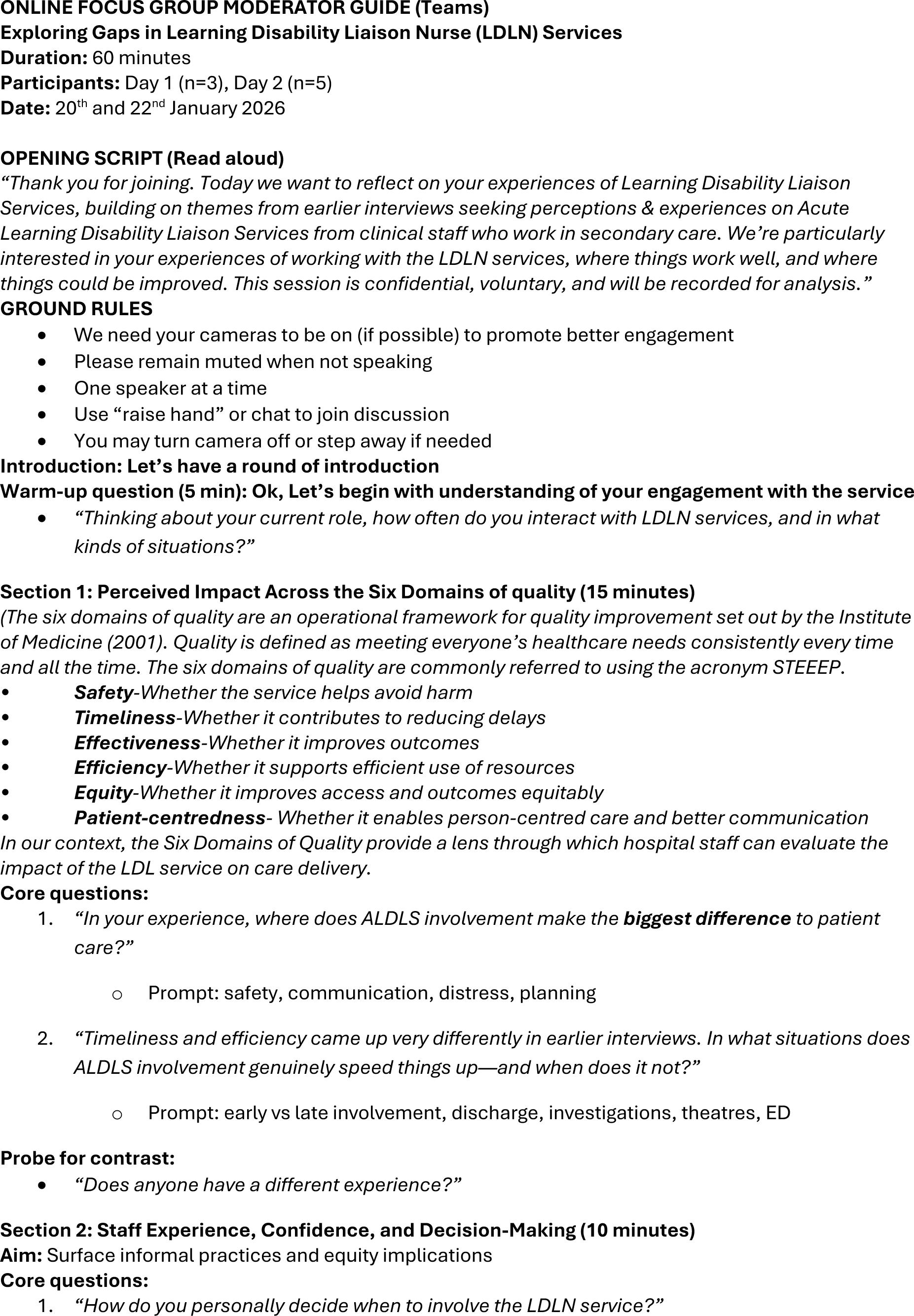
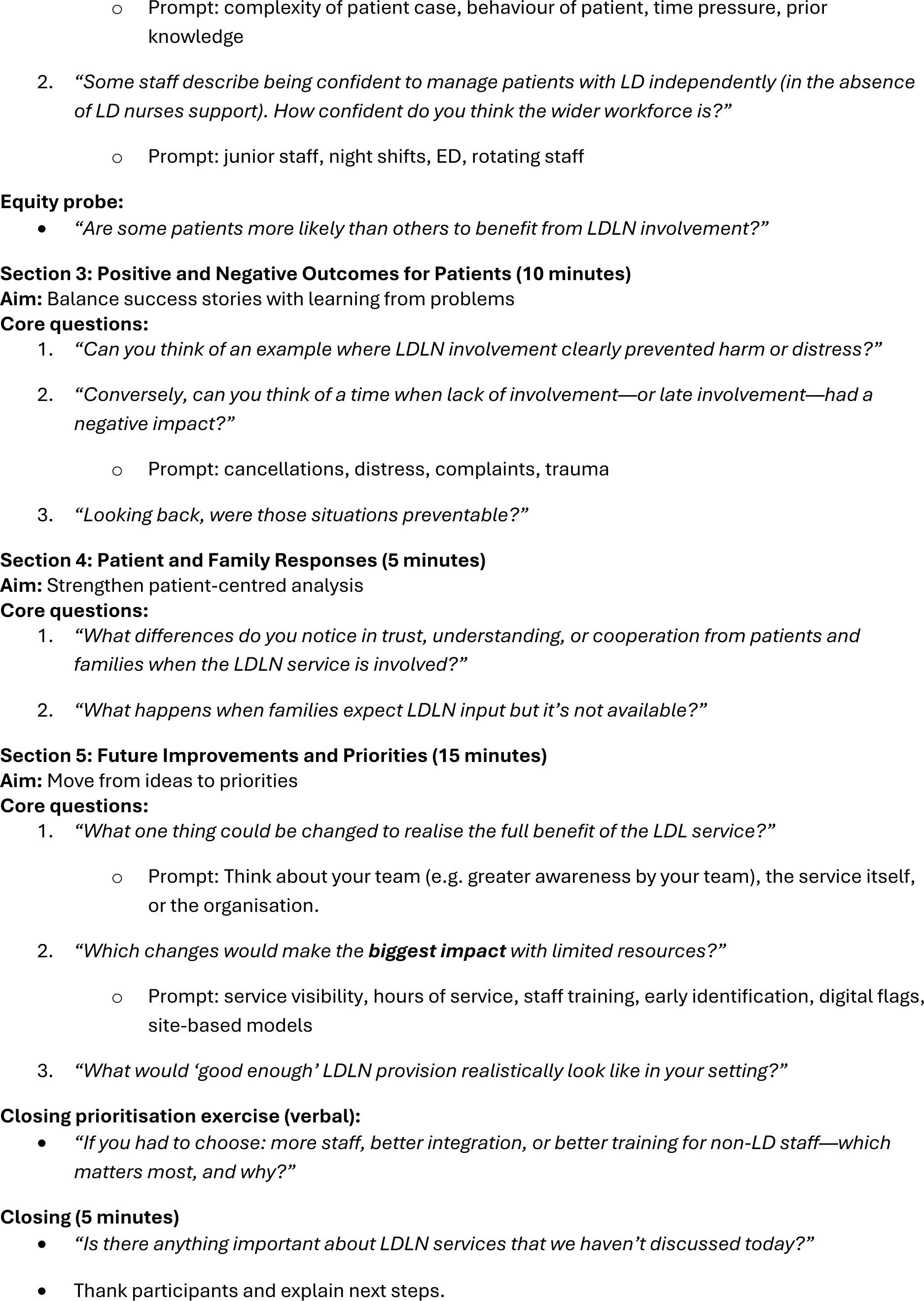

## ABBREVIATIONS

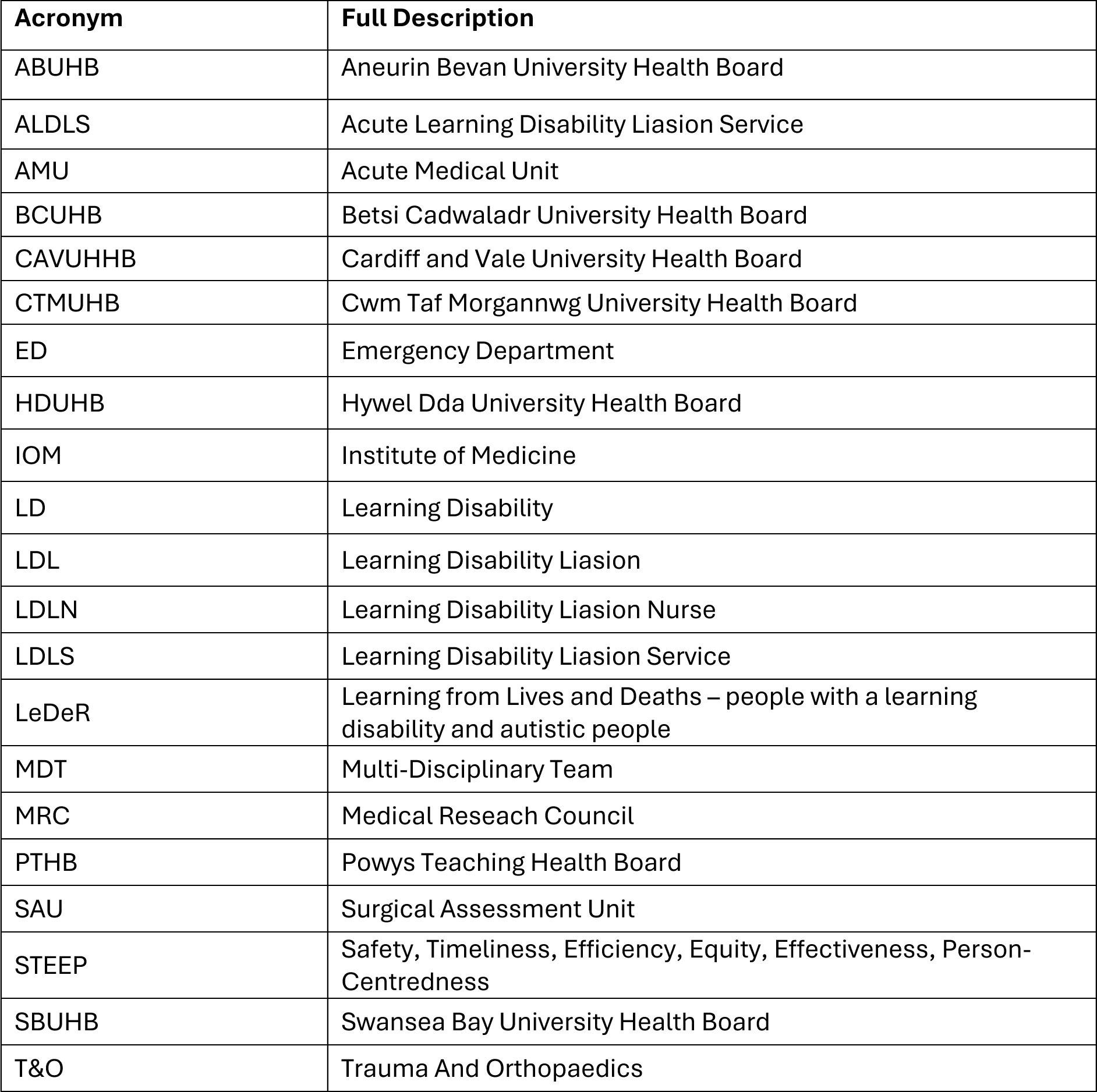

